# Cardiac MRI measures as surrogate outcome for heart failure and atrial fibrillation: a Mendelian randomization analysis

**DOI:** 10.1101/2022.07.27.22278120

**Authors:** A F Schmidt, C Finan, J van Setten, E Puyol-Anton, B Ruijsink, M Bourfiss, A I Alasiri, B K Velthuis, F W Asselbergs, A S J M te Riele

**Author notes:** **Corresponding author:** Amand Floriaan Schmidt, PhD.

## Abstract

**Background:** drug development and disease prevention of heart failure (HF) and atrial fibrillation (AF) are impeded by a lack of robust early-stage surrogates. We determined to what extent cardiac magnetic resonance (CMR) measurements act as surrogates for the development of HF or AF in healthy individuals.

**Methods:** Genetic data was sourced on the association with 22 atrial and ventricular CMR measurements. Mendelian randomization was used to determine CMR associations with atrial fibrillation (AF), heart failure (HF), non-ischemic cardiomyopathy (CMP), and dilated cardiomyopathy (DCM). Additionally, for the CMR surrogates of AF and HF, we explored their association with non-cardiac traits.

**Results:** In total we found that 10 CMR measures were associated with the development of HF, 8 with development of non-ischemic CMP, 5 with DCM, and 11 with AF. Left-ventricular (LV) ejection fraction (EF), and LV end diastolic volume (EDV) were associated with all 4 cardiac outcomes. Increased LV-MVR (mass to volume ratio) affected HF (odds ratio (OR) 0.83, 95%CI 0.79; 0.88), DCM (OR 0.26, 95%CI 0.20; 0.34), non-ischemic CMP (OR 0.44 95%CI, 0.35; 0.57). We were able to identify 9 CMR surrogates for HF and AF (including LV-MVR, biventricular EDV, right-ventricular EF, and left-atrial maximum volume) which associated with non-cardiac traits such as blood pressure, cardioembolic stroke, diabetes, and late-onset Alzheimer’s disease.

**Conclusion:** CMR measurements may act as surrogate endpoints for the development of HF (including non-ischemic CMP and DCM) or AF. Additionally, we show that changes in cardiac function and structure measured through CMR, may affect diseases of other organs leading to diabetes and late-onset Alzheimer’s disease.

## Introduction

Heart failure (HF) and atrial fibrillation (AF) are major cardiac diseases that cause considerable burden in terms of health and economic costs, as well as mortality^1–3^. HF is a clinical syndrome secondary to dysfunction of the right ventricle (RV) or left ventricle (LV), while AF is defined by uncoordinated electrical activation and consequently ineffective contraction of the atria. Both diseases are intricately related and while the causative relationship between the two conditions has not been fully determined, it is clear these two syndromes frequently co-occur^4^.

Despite recent advances in medicines, for example offered by sodium-glucose co-transporter-2 inhibitors, drug development for cardiac disease suffers from high failure rates, often occurring during costly late-stage clinical testing^5–7^. Unlike with the cholesterol content on low-density lipoprotein for coronary heart disease, drug development for AF and HF is impeded by a lack of robust early-stage surrogates (or intermediates) for cardiac disease.

Cardiac magnetic resonance (CMR) imaging is the gold standard for quantification of atrial and ventricular function and morphology, and has become an integral diagnostic modality for cardiac diseases. It is however unclear to what extent CMR measurements act as surrogates for the development of cardiac disease in otherwise healthy individuals.

Both HF and AF are associated with multimorbidity including non-cardiac diseases, such as stroke, chronic kidney disease (CKD), diabetes mellitus, and neurological diseases such as Alzheimer’s disease. Because HF and AF are clinical manifestations of underlying changes in cardiac function and structure, patients with similar diagnoses may vary considerably in underlying pathophysiology and disease progression. Unlike HF or AF diagnoses, CMR measurements directly reflect cardiac physiology, and therefore provide an opportunity to explore the effects changes in cardiac function and structure may elicit in other organs.

Recently, CMR measurements of thousands of subjects have been linked to genetic data and analysed through genome-wide analysis studies (GWAS). Aggregate data from GWAS, consisting of variant-specific point estimates and standard errors, can be used in Mendelian randomization analyses to ascertain the causal effects a CMR trait may have on disease. In the current manuscript, we leveraged data from three recent GWAS of CMR measurements of atrial and ventricular structure and function^8^, LV trabeculation morphology^9^, and left atrial (LA) volume^10^, jointly consisting of 22 measurements conducted in over 35,000 UK biobank (UKB) participants. These data were used to conduct Mendelian randomization analyses of the association between CMR traits and cardiac events, including HF, dilated cardiomyopathy (DCM), non-ischemic cardiomyopathy (CMP), and AF. Subsequently, we explored the association of CMR proxies for HF or AF with 20 clinically relevant non-cardiac traits.

## Methods

### Genetic data on CMR and cardiac traits

We leveraged aggregate data (i.e., point estimates and standard errors) from 3 GWAS of deep-learning derived CMR measurements conducted using UKB participants; please see the specific study references for details on the derivation methods. Ahlberg *et al*.^10^ provided measurements on LA volume (LA-V (max) and LA-V (min)), LA total emptying fraction (LA-TF), LA active emptying fraction (LA-AF), and passive emptying fraction (LA-PF) from 35,658 subjects. Genetic data on LV trabecular morphology (LV-TM), measured as a fractal dimension ratio, was available from Meyer *et al*.^9^ on 18,096 subjects. Schmidt *et al*^8^. provided (n: 36,548) data on LV and right-ventricular (RV) ejection fraction (EF), stroke volume (SV), peak filling rate (PFR), peak ejection rate (PER), end-diastolic or end-systolic volumes (EDV, ESV), LV end-diastolic mass (LV-EDM), the LV mass to volume ratio (LV-MVR), and biatrial PFR. All three GWAS excluded subjects with pre-existing cardiac conditions such as AF, HF, cardiomyopathies, myocardial infarction, or congenital heart disease.

GWAS data was sourced on the following cardiac outcomes: HF (52,496 cases)^11^, non-ischemic CMP (1,816 cases)^12^, DCM (2,719 cases)^13^, and AF (60,620 cases)^14^. The following 20 traits were used in the non-cardiac phenome-wide scan: stroke (subtypes), venous thromboembolism (VTE), abdominal aortic aneurysm (AAA), systolic/diastolic blood pressure (SBP/DBP), body mass index (BMI), diabetes (T2DM), glycated haemoglobin, c-reactive protein (CRP), forced expiratory volume (FEV1), forced vital capacity (FVC), chronic kidney disease (CKD), estimated glomerular filtration rate (eGFR), Alzheimer’s disease (AD), late-onset AD, and late Lewy body dementia; please see the data availability section for more detail.

### Mendelian randomization analysis

Genetic instruments were selected from throughout the genome using an F-statistic > 24 and a minor allele frequency (MAF) of at least 0.01. Variants were clumped to a linkage disequilibrium (LD) R-squared threshold of 0.30, with residual LD modelled using a generalized least square (GLS) solution^15^ and a reference panel from a random sample of 5,000 of white British ancestry UKB participants; this following the source GWAS data excluding non-European ancestries to prevent bias through population stratification.

Mendelian randomization was conducted using the GLS implementation of the inverse-variance weighted (IVW) estimator, as well as with an Egger correction to protect against horizontal pleiotropy^16^. To further minimize the potential influence of horizontal pleiotropy, we excluded variants with large leverage or outlier statistics, and used the Q-statistic to identify possible remaining violations^17^. Finally, a model selection framework was applied to select the most appropriate estimator (IVW or MR-Egger) for each individual exposure-outcome relation^17,18^.

Where appropriate, results were presented as odds ratio (OR, for binary traits) with 95% confidence interval (95%CI) or mean difference (MD, for continuous traits). Associations with cardiac outcomes were declared significant using the standard alpha of 0.05, with a multiplicity corrected alpha of 1.25×10^−2^ (correcting for the 20 non-cardiac traits) applied to the more exploratory phewas analysis. Under the null-hypothesis (i.e., where all results are false positives) p-values follow a standard uniform distribution. Hence, to identify CMR associations driven by multiplicity we performed Kolmogorov-Smirnov “KS”-tests^19^ evaluating the agreement of the empirical p-value distribution with the standard uniform distribution.

## Results

### Biventricular and atrial CMR associations with incident cardiac outcomes

Sourcing CMR measurements in people without pre-existing cardiac conditions we employed Mendelian randomization to determine their association with the development of cardiac events. Higher EF of both ventricles was associated with decreased risk of HF, non-ischemic CMP, and DCM; Figure 1, Supplementary Table 1. Higher RV-PFR increased the risk of AF (OR 2.12, 95%CI 1.15; 3.90). Higher EDV and ESV of both ventricles protected against AF (excluding RV-ESV), while increasing the risk of HF (excluding RV-EDV), non-ischemic CMP(excluding RV-ESV), and DCM (excluding RV-EDV); Figure 1. Higher LV-MVR decreased the risks of HF (OR 0.83, 95%CI 0.79; 0.88), non-ischemic CMP(OR 0.44, 95%CI 0.35; 0.57), and DCM (OR 0.26, 95%CI 0.20; 0.34). From the atrial traits presented in Figure 2, higher LA-V (min) increased HF risk (OR 1.12 95%CI 1.00; 1.26), and AF risk (OR 1.18, 95%CI 1.00; 1.40). Out of the atrial emptying measures AF risk was decreased by higher LA-TF (OR 0.83, 95%CI 0.72; 0.97) and higher LA-AF (OR 0.77, 95%CI 0.70; 0.85), with the latter also decreasing HF risk (OR 0.85, 95%CI 0.78; 0.92).

**Figure 1.**
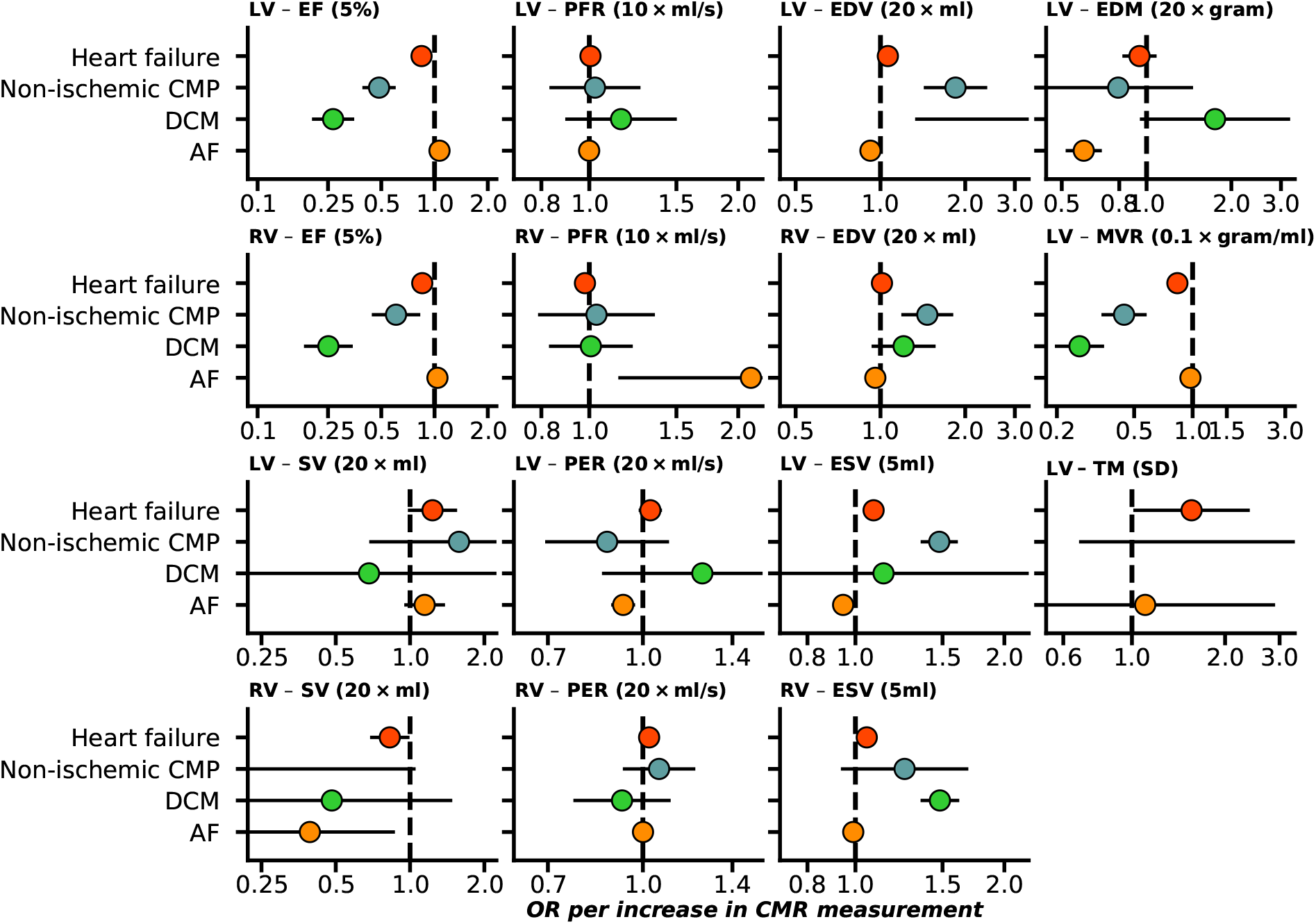
Forest plot of Mendelian randomization estimates of biventricular CMR associations with the onset of HF and AF. n.b. Point estimates reflect odds ratios (OR) with 95% confidence intervals presented as horizontal line segments. LV: left-ventricle, RV: right-ventricle, EF: ejection fraction, SV: stroke volume, PFR: peak filling rate, PER: peak ejection rate, EDV/ESV: diastolic or systolic volumes, EDM: end diastolic mass, MVR: mass to volume ratio, TM: trabecular morphology. Outcome data was available on heart failure (52,496 cases), DCM (dilated cardiomyopathy, 2,719 cases), non-ischemic CMP(1,816 cases), and AF (atrial fibrillation, 60,620 cases).

**Figure 2.**
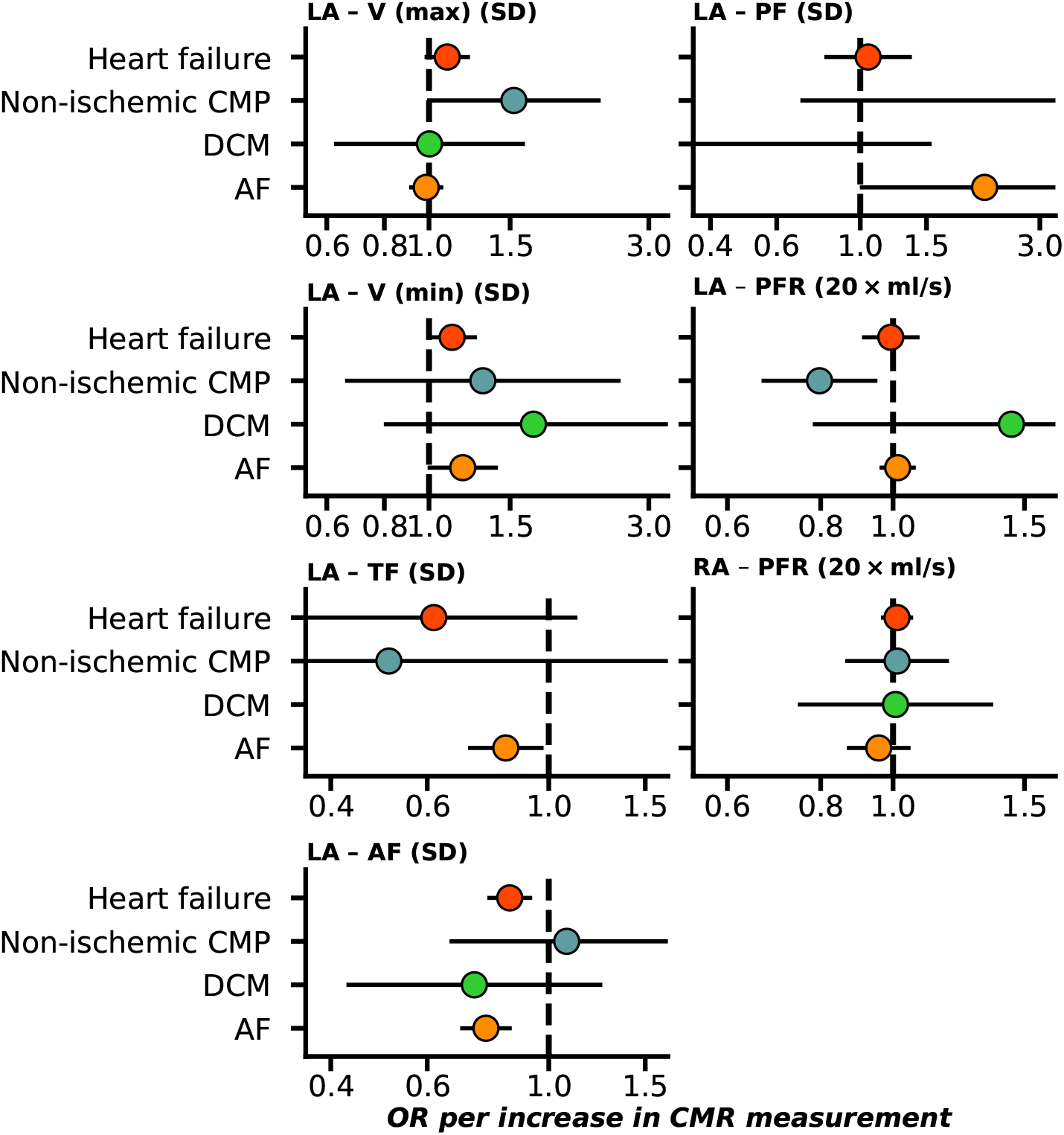
Forest plot of Mendelian randomization estimates of atrial CMR associations with the onset of HF and AF. n.b. Point estimates reflect odds ratios (OR) with 95% confidence intervals presented as horizontal line segments. RA: right-atrial, LA: left-atrial, V (max): maximum volume, V (min): minimum volume, TF: total emptying fraction, AF: active emptying fraction, PF: passive emptying fraction, PFR: peak filling rate. Outcome data was available on heart failure (52,496 cases), DCM (dilated cardiomyopathy, 2,719 cases), non-ischemic CMP(1,816 cases), and AF (atrial fibrillation, 60,620 cases).

In general, we found that AF occurrence was affected by changes in atrial function, as well as changes in structure and function of LV and RV (Figures 1-3), including RV-PFR, LV-EDV, LV-EDM, LV-MVR, LA-TF, and LA-AF. We similarly observed the importance of changes in biventricular structure and function in the occurrence of HF and cardiomyopathies, where left-atrial functioning was important as well (Figures 1-3).

**Figure 3.**
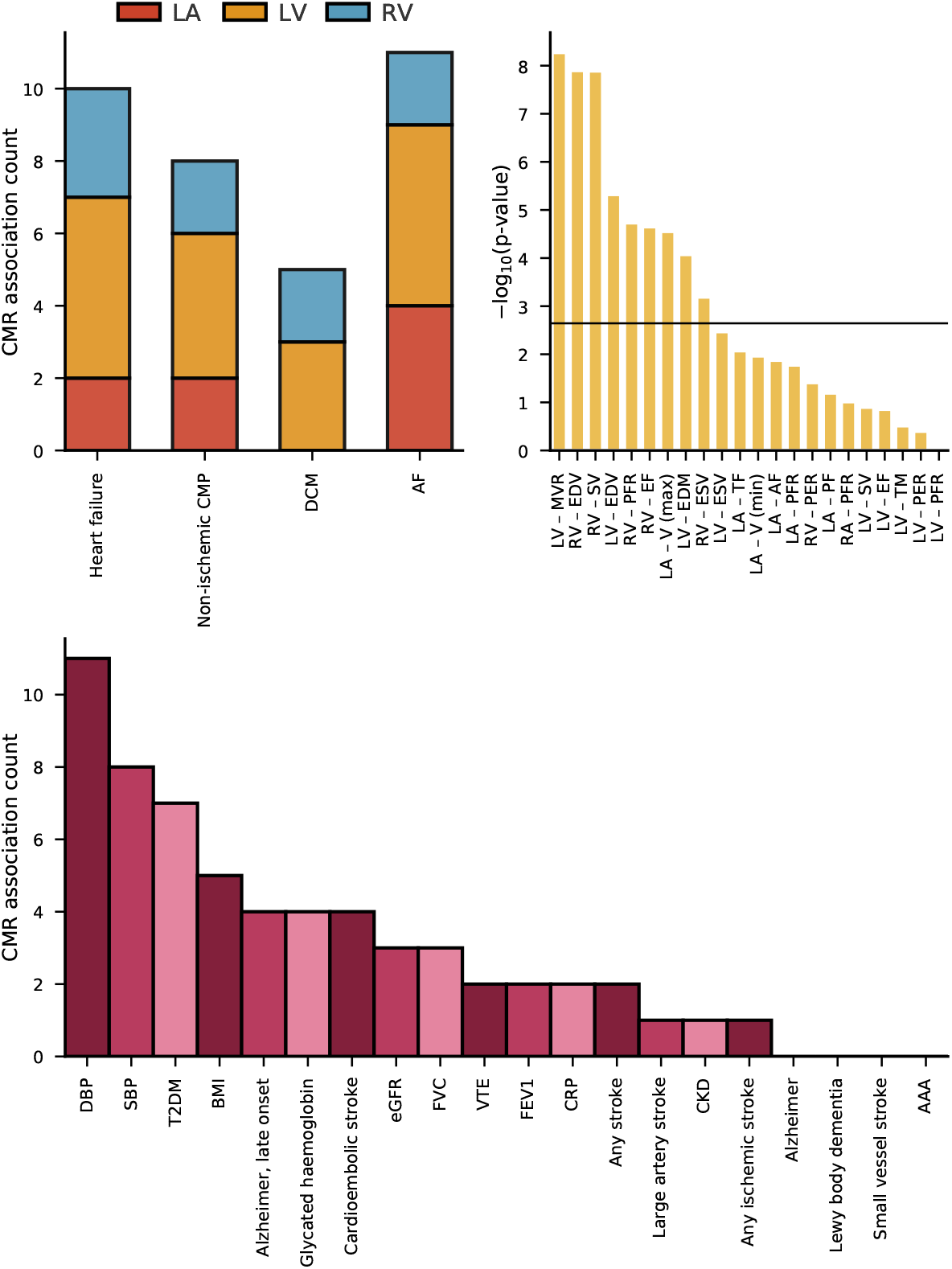
CMR association counts with cardiac and non-cardiac traits, and Kolmogorov-Smirnov test results for multiplicity. n.b The **top left** bar chart represents the counts of significant Mendelian randomization CMR effects grouped by chamber. The **top right** bar chart represent the -log_10_(p-value) of Kolmogorov-Smirnov tests, were a significant result indicates the phewas results are unlikely driven by multiple testing. The horizontal line indicates the significance threshold of 0.05/20. The **bottom** bar chart represents the counts of significant Mendelian randomization CMR effects on the considered phewas traits. The following abbreviations were used, LV: left-ventricle, RV: right-ventricle, RA: right-atrial, LA: right-atrial, HCM: hypertrophic cardiomyopathy, DCM: dilated cardiomyopathy, AF: atrial fibrillation, T2DM: type 2 diabetes, CKD: chronic kidney disease, VTE: venous thromboembolism, AAA: abdominal aortic aneurysm, SBP/DBP: systolic/diastolic blood pressure, BMI: body mass index, CRP: c-reactive protein, FVC: forced vital capacity, FEV1: forced expiratory volume, eGFR: estimated glomerular filtration rate, HbA1c: glycated haemoglobin.

### Associations of cardiac function and structure with non-cardiac traits

We next explored whether changes in cardiac function and structure could be associated with non-cardiac traits. We found that CMR traits were frequently associated with blood pressure, incidence type 2 diabetes (T2DM), BMI, late onset (after an age of 65 years) Alzheimer’s disease (AD), and cardioembolic stroke (Figure 3, Supplementary Table 2).

Prioritizing results that were unlikely driven by multiple testing (Figures 3-4), we identified 9 CMR surrogates for HF and/or AF that were associated with non-cardiac traits: LV-MVR, LV-EDM, biventricular EDV, RV-SV, RV-ESV, RV-EF, RV-PFR, and LV-A (max). For example, we observed that all the aforementioned CMR measures of cardiac structure or function associated with blood pressure (Figure 4). We additionally observed that increased LV-MVR (OR 0.63, 95%CI 0.55; 0.73) decreased the risk of cardioembolic stroke. LV-EDV and LV-EDM were associated with a decreased risk of late-onset AD (Figure 4): OR 0.82 (95%CI 0.75; 0.90) and OR 0.68 (95%CI 0.55; 0.84), respectively. BMI was affected by changes in functional parameters, specifically by RV-EF, RV-EDV and RV-ESV, and T2DM risk was driven by RV-SV, RV-PFR, RV-EDV, LV-EDM, LV-MVR, and LV-V (max). LV-MVR was a particularly important measure, associating with most stroke types, VTE, SBP, DPB, T2DM, CRP, and eGFR; Figure 4.

**Figure 4.**
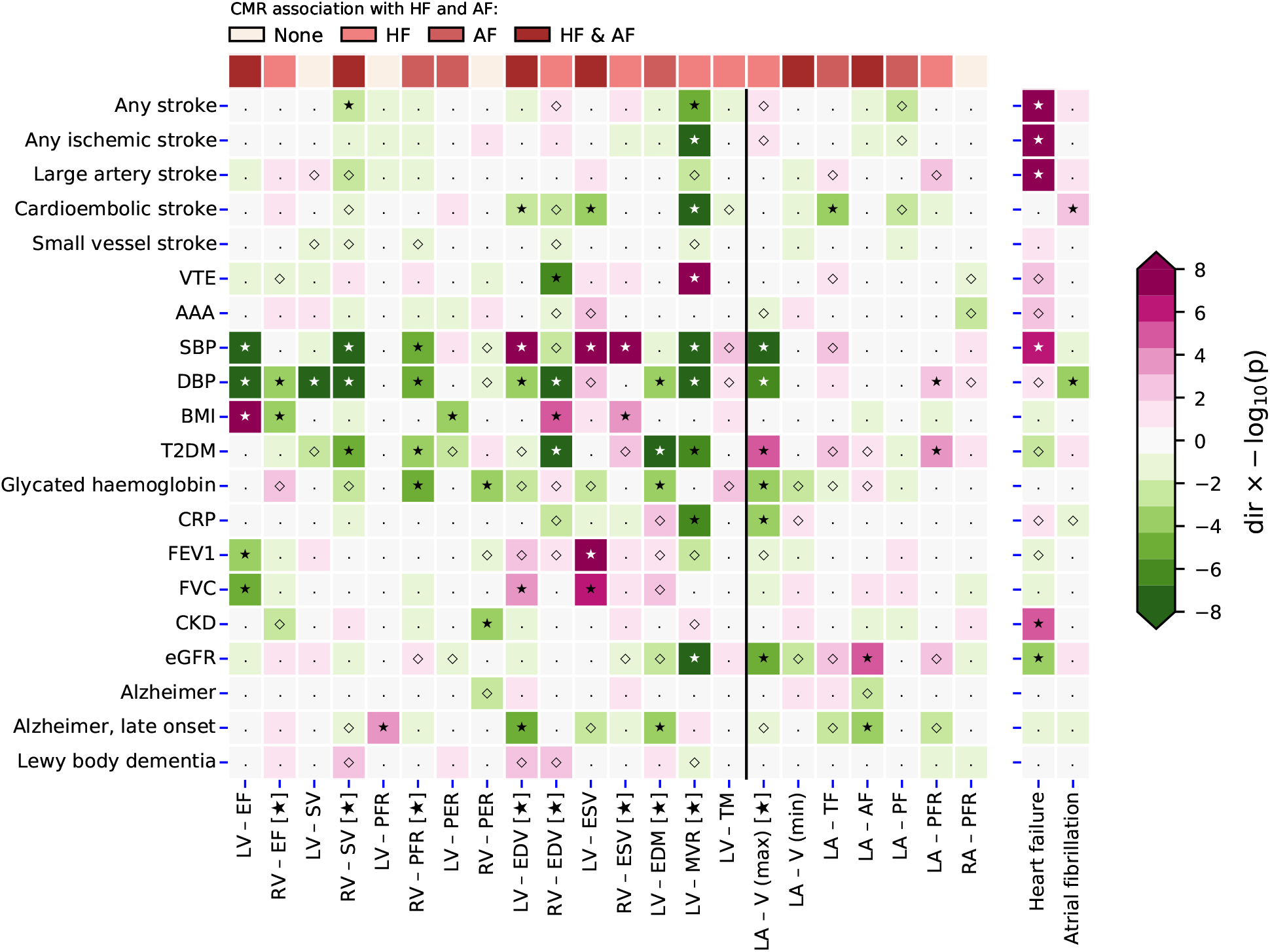
A targeted Mendelian randomization phenome-wide scan comparing the effect of changes in cardiac function and structure to that of a HF or AF diagnosis. N.b. P-values passing the 0.05 threshold are indicated by an open diamond with stars indicating results passing a threshold of 1.25×10^−2^. Cells were coloured by effect direction times -log_10_(p-value); where p-values were truncated at 8 for display purposes. CMR measurements that passed the Kolmogorov-Smirnov test for multiplicity are indicated with a star, with results of non-indicated CMR measurements more likely driven by multiplicity. The following abbreviations were used, LV: left-ventricle, RV: right-ventricle, RA: right-atrial, LA: right-atrial, EF: ejection fraction, SV: stroke volume, PFR: peak filling rate, PER: peak ejection rate, EDV/ESV: diastolic or systolic volumes, EDM: end diastolic mass, MVR: mass to volume ratio, TM: trabecular morphology V (max): maximum volume, V (min): minimum volume, TF: total emptying fraction, AF: active emptying fraction, PF: passive emptying fraction, AF: atrial fibrillation, T2DM: type 2 diabetes, CKD: chronic kidney disease, VTE: venous thromboembolism, AAA: abdominal aortic aneurysm, SBP/DBP: systolic/diastolic blood pressure, BMI: body mass index, CRP: c-reactive protein, FVC: forced vital capacity, FEV1: forced expiratory volume, eGFR: estimated glomerular filtration rate, HbA1c: glycated haemoglobin.

### Comparison to HF and AF effects on non-cardiac traits

Next, as comparison we leveraged genetic instruments with a clinical diagnosis of HF or AF, and performed Mendelian randomization to determine the causal effects HF or AF had on non-cardiac traits. HF increased the risk of any stroke, any ischemic stroke, as well as large artery stroke, SBP and chronic kidney disease (Figure 4, Supplementary Table 3). AF diagnosis increased the risk of cardioembolic stroke (OR 2.13 95%CI 1.35; 3.34) and associated with DBP.

## Discussion

In the current manuscript we employed Mendelian randomization combined with CMR measurements in participants without pre-existing cardiac disease and identified surrogate outcomes for the onset of HF (52,496 cases) and AF (60,620 cases). We show that biventricular EF, EDV, ESV, LV-MVR, and LA-AF are associated with a decreased risk of *de novo* development of HF, non-ischemic CMP, DCM, and/or AF. Importantly, we found that the development of HF or AF is not exclusively driven by any single ventricular or atrial measurement, but is determined by combinations of changes in atrial and ventricular function and structure.

In total, we identified 10 CMR measures associated with the development of HF, 8 with development of non-ischemic CMP, 5 with DCM, and 11 with AF. This indicates that CMR measurements in healthy individuals can be used to monitor disease occurrence and help identify high-risk patients in need of preventative measures. Additionally, our findings imply that CMR measurement might be used as surrogate endpoints in early clinical studies, which can assist in prioritizing compounds for confirmatory outcome trials.

We explored the phenotypic effects that changes in cardiac function and structure may have on non-cardiac traits (Figure 3-4), identifying 9 CMR surrogates (5 RV, 3 LV and 1 LA CMR measurements) for HF or AF that also associated with non-cardiac traits. All 9 CMR measurements were associated with SBP and DBP, confirming the well-established relation between HF and blood pressure^20^. Additionally, we observed that increased LV-EDV and LV-MVR were associated with a decreased likelihood of cardioembolic stroke, where LV-EDV also decreased the onset of AF and LV-MVR decreased the risk of HF and DCM, which are known risk factors for cardioembolic stroke^21,22^. Changes in RV function such as indicated by SV, EDV and PFR were associated with T2DM, providing support for shared aetiology with diabetic cardiomyopathy^23^. Increased LV-EDV and LV-EDM were associated with lower blood pressure as well as decreased risk of late-onset AD, suggesting a blood pressure mediated AD effect of LV changes in function and structure^24^.

Interestingly, LV-EF was not strongly associated with the development of non-cardiac disease. Instead, we observed a strong association of LV-MVR with over 10 non-cardiac traits (Figure 4), including stroke subtypes, VTE, and T2DM. This suggests that while LV-EF has important diagnostic implications for HF, a broader consideration of CMR measurements might provide further information relevant for risk mitigation of diseases often co-occurring in people at high risk of developing HF or AF. This is further highlighted by our finding that 5 of the 9 CMR measurement associated with non-cardiac traits were RV, supporting the need to for a more holistic consideration changes in cardiac function and structure may have on disease risk.

The study has a number of limitations that deserve consideration. First, while we sourced genetic associations with CMR measurement taken from subjects without pre-existing cardiac conditions, a proportion of subjects may have had undiagnosed disease. The UKB however represents a relatively healthy subset of the UK population, likely minimizing the number of individuals with latent disease. Second, our choice of CMR measurement was limited by the publicly available data, for example preventing us from exploring the association between ratio of measure (such as PEF/EDV or PFR/EDV) not available in the original results. Third, while Mendelian randomization is robust against bias through reverse causality and confounding bias, it critically assumes the absence of horizontal pleiotropy, where the genetic variant only affects the outcome through its association with the CMR measurement. In the current analysis, we performed automatic model selection to decide between an IVW or more robust MR-egger models, and additionally removed potentially pleiotropic variants through the identification and removal of outliers and high leverage points. Fourth, due to its protection against reverse causation, Mendelian randomization results are naturally imbued with a clear directionality of association. In the current analyses this means that the observed Mendelian randomization estimates proxy the effects underlying changes in cardiac function or morphology may have on the considered outcomes. This does not however preclude more complex bi-directional or synergistic disease pathways. For example, our observation that CMR measures that decrease HF risk also decrease T2DM risk, is in line with previous observational analyses suggesting that people with T2DM are at risk of HF^27^, and implies either a bi-directional effects between HF and T2DM, or that the CMR effect on HF is partially mediated by changes in glucose metabolism. Finally, the conducted Mendelian randomization analyses implicitly assess a linear trend between CMR and outcome. In the presence of non-linearity, the presented Mendelian randomization estimates represent a population average effect which may not necessarily apply to any single individual, but often offers a reasonably approximation. While non-linear Mendelian randomization methods have been developed^25,26^, these require access to individual participant data which, even for UKB sized data, only offer a fraction of the disease cases we have been able to leverage here.

In conclusion, we have identified biventricular and left-atrial CMR measurements that may act as surrogate endpoints for future cardiac events, including heart failure, cardiomyopathies, and atrial fibrillation. We additionally show that changes in cardiac function and structure may affect other organs, resulting in diseases such as diabetes and late-onset Alzheimer’s.

## Data Availability

Genetic data for the phewas of non-cardiac traits was sourced for stroke (subtypes) from MEGASTROKE (http://www.megastroke.org/index.html), venous thromboembolism and abdominal aortic aneurysm from (https://www.globalbiobankmeta.org/), blood pressure from Evangelou et.al. (https://www.nature.com/articles/s41588-018-0205-x), glycemic traits, and lung function measurement were sourced from (http://www.nealelab.is/uk-biobank); type 2 diabetes from DIAGRAM (http://diagram-consortium.org/index.html); BMI from GAINT (https://portals.broadinstitute.org/collaboration/giant/index.php/GIANT_consortium_data_files); CRP from (https://www.ebi.ac.uk/gwas/publications/30388399); the CKDGen consortium provided GWAS associations on estimated glomerular filtration rate (eGFR), and chronic kidney disease (http://ckdgen.imbi.uni-freiburg.de/); Alzheimer's disease data was sourced from Jansen et.al. (https://ctg.cncr.nl/software/summary_statistics) and Kunkle et.al. (https://www.nature.com/articles/s41588-019-0358-2); Lewy body dementia from (https://www.ebi.ac.uk/gwas/publications/33589841);

## Data availability

Genetic data for the phewas of non-cardiac traits was sourced for stroke (subtypes) from MEGASTROKE (http://www.megastroke.org/index.html), venous thromboembolism and abdominal aortic aneurysm from (https://www.globalbiobankmeta.org/), blood pressure from Evangelou *et*.*al*. (https://www.nature.com/articles/s41588-018-0205-x), glycemic traits, and lung function measurement were sourced from (http://www.nealelab.is/uk-biobank); type 2 diabetes from DIAGRAM (http://diagram-consortium.org/index.html); BMI from GAINT (https://portals.broadinstitute.org/collaboration/giant/index.php/GIANT_consortium_data_files); CRP from (https://www.ebi.ac.uk/gwas/publications/30388399); the CKDGen consortium provided GWAS associations on estimated glomerular filtration rate (eGFR), and chronic kidney disease (http://ckdgen.imbi.uni-freiburg.de/); Alzheimer’s disease data was sourced from Jansen *et*.*al*. (https://ctg.cncr.nl/software/summary_statistics) and Kunkle *et*.*al*. (https://www.nature.com/articles/s41588-019-0358-2); Lewy body dementia from (https://www.ebi.ac.uk/gwas/publications/33589841);

## Code availability

Analyses were conducted using Python v3.7.10 (for GNU Linux), Pandas v1.3.5, Numpy v1.20.3, and Matplotlib 3.3.2. Scripts and data necessary to generate the illustrations have been deposited: XX.

## Author’s contributions

AFS, FWA, ASJM contributed to the idea and design of the study. AFS performed the analyses. AFS, and ASJM drafted the manuscript. All authors provided critical input on the analyses and the drafted manuscript.

## Guarantor

AFS performed the presented analyses. AFS had full access to all the data in the study and takes responsibility for the integrity of the data and the accuracy of the data analysis.

## Acknowledgement

This research has been conducted using the UK Biobank Resource under Application Numbers 24711, 12113, and 44972. The authors are grateful to UK Biobank participants. UK Biobank was established by the Wellcome Trust medical charity, Medical Research Council, Department of Health, Scottish Government, and the Northwest Regional Development Agency. It has also had funding from the Welsh Assembly Government and the British Heart Foundation.

## Prior postings and presentations

A preprint version of this manuscript has been deposited on medrxiv:.

