## Supplementary File 1 for "Cardiac MRI measures as surrogate outcome for heart failure and atrial fibrillation: a Mendelian randomization analysis"

A F Schmidt *et al.*

#### Contents

**Tables** **2**

### Tables

**Supplementary Table 1:** Mendelian randomization results on the effects of CMR measured changes in cardiac function and structure have on incident cardiac diagnoses.

| CMR measure | Units | Cardiac diagnosis | OR (95%CI) | P-value | Q p-value | No. variants | MR model |
|---|---|---|---|---|---|---|---|
| LV – EF | 5 % | Heart failure | 0.84 (0.81; 0.89) | $1.4\times10^{-12}$ | 0.003 | 43 | IVW |
| | | DCM | 0.27 (0.21; 0.34) | $1.0\times10^{-100}$ | <0.001 | 40 | IVW |
| | | Non-ischemic CMP | 0.49 (0.40; 0.59) | $3.9\times10^{-13}$ | 0.007 | 44 | IVW |
| | | AF | 1.07 (1.01; 1.12) | $1.1\times10^{-2}$ | <0.001 | 40 | IVW |
| RV – EF | 5 % | Heart failure | 0.85 (0.81; 0.90) | $8.0\times10^{-9}$ | 0.004 | 39 | IVW |
| | | DCM | 0.25 (0.19; 0.34) | $1.0\times10^{-100}$ | <0.001 | 39 | IVW |
| | | Non-ischemic CMP | 0.61 (0.45; 0.81) | $8.2\times10^{-4}$ | 0.051 | 40 | IVW |
| | | AF | 1.04 (0.99; 1.09) | $1.5\times10^{-1}$ | <0.001 | 41 | IVW |
| LV – SV | 20 ml | Heart failure | 1.23 (1.00; 1.53) | $5.5\times10^{-2}$ | 0.133 | 8 | IVW |
| | | DCM | 0.68 (0.18; 2.56) | $5.7\times10^{-1}$ | 0.078 | 8 | IVW |
| | | Non-ischemic CMP | 1.58 (0.69; 3.59) | $2.8\times10^{-1}$ | 0.800 | 8 | IVW |
| | | AF | 1.15 (0.96; 1.36) | $1.3\times10^{-1}$ | 0.714 | 8 | IVW |
| RV – SV | 20 ml | Heart failure | 0.83 (0.70; 0.98) | $2.8\times10^{-2}$ | <0.001 | 11 | IVW |
| | | DCM | 0.48 (0.16; 1.46) | $2.0\times10^{-1}$ | 0.063 | 11 | IVW |
| | | Non-ischemic CMP | 0.08 (0.01; 1.04) | $5.3\times10^{-2}$ | 0.381 | 11 | MR Egger |
| | | AF | 0.39 (0.18; 0.85) | $1.9\times10^{-2}$ | 0.066 | 9 | MR Egger |
| LV – PFR | 10 ml/s | Heart failure | 1.01 (0.98; 1.03) | $7.1\times10^{-1}$ | 0.465 | 2 | IVW |
| | | DCM | 1.16 (0.90; 1.49) | $2.5\times10^{-1}$ | 0.154 | 2 | IVW |
| | | Non-ischemic CMP | 1.03 (0.84; 1.26) | $8.0\times10^{-1}$ | 0.124 | 2 | IVW |
| | | AF | 1.00 (0.96; 1.04) | $9.7\times10^{-1}$ | 0.323 | 2 | IVW |
| RV – PFR | 10 ml/s | Heart failure | 0.98 (0.95; 1.01) | $1.9\times10^{-1}$ | 0.500 | 2 | IVW |
| | | DCM | 1.01 (0.84; 1.21) | $9.4\times10^{-1}$ | 0.385 | 2 | IVW |
| | | Non-ischemic CMP | 1.03 (0.79; 1.35) | $8.0\times10^{-1}$ | 0.069 | 2 | IVW |
| | | AF | 2.12 (1.15; 3.90) | $1.6\times10^{-2}$ | None | 2 | MR Egger |
| LV – PER | 20 ml/s | Heart failure | 1.03 (0.99; 1.07) | $1.4\times10^{-1}$ | 0.480 | 4 | IVW |
| | | DCM | 1.25 (0.86; 1.81) | $2.4\times10^{-1}$ | 0.148 | 3 | IVW |
| | | Non-ischemic CMP | 0.87 (0.70; 1.10) | $2.4\times10^{-1}$ | 0.454 | 3 | IVW |
| | | AF | 0.93 (0.89; 0.97) | $1.7\times10^{-4}$ | 0.785 | 4 | IVW |
| RV – PER | 20 ml/s | Heart failure | 1.02 (0.99; 1.06) | $1.3\times10^{-1}$ | 0.665 | 8 | IVW |
| | | DCM | 0.92 (0.78; 1.10) | $3.8\times10^{-1}$ | 0.600 | 8 | IVW |
| | | Non-ischemic CMP | 1.06 (0.93; 1.21) | $3.6\times10^{-1}$ | 0.555 | 8 | IVW |
| | | AF | 1.00 (0.97; 1.03) | $9.7\times10^{-1}$ | 0.755 | 9 | IVW |
| LV – EDV | 20 ml | Heart failure | 1.06 (1.00; 1.13) | $4.4\times10^{-2}$ | 0.003 | 26 | IVW |
| | | DCM | 5.57 (1.35; 23.07) | $1.8\times10^{-2}$ | <0.001 | 27 | MR Egger |
| | | Non-ischemic CMP | 1.85 (1.44; 2.36) | $1.0\times10^{-6}$ | 0.005 | 25 | IVW |
| | | AF | 0.92 (0.87; 0.98) | $5.5\times10^{-3}$ | 0.032 | 22 | IVW |
| RV – EDV | 20 ml | Heart failure | 1.01 (0.96; 1.07) | $6.3\times10^{-1}$ | <0.001 | 36 | IVW |
| | | DCM | 1.21 (0.94; 1.55) | $1.3\times10^{-1}$ | <0.001 | 38 | IVW |
| | | Non-ischemic CMP | 1.47 (1.20; 1.79) | $1.6\times10^{-4}$ | 0.013 | 37 | IVW |
| | | AF | 0.96 (0.92; 1.00) | $5.2\times10^{-2}$ | <0.001 | 37 | IVW |
| LV – ESV | 5 ml | Heart failure | 1.09 (1.07; 1.11) | $1.0\times10^{-100}$ | <0.001 | 58 | IVW |
| | | DCM | 1.14 (0.43; 3.02) | $7.9\times10^{-1}$ | <0.001 | 31 | MR Egger |
| | | Non-ischemic CMP | 1.48 (1.37; 1.60) | $1.0\times10^{-100}$ | <0.001 | 54 | IVW |
| | | AF | 0.94 (0.93; 0.96) | $5.7\times10^{-12}$ | <0.001 | 52 | IVW |
| RV – ESV | 5 ml | Heart failure | 1.05 (1.04; 1.07) | $1.9\times10^{-10}$ | <0.001 | 52 | IVW |
| | | DCM | 1.48 (1.36; 1.61) | $1.0\times10^{-100}$ | <0.001 | 55 | IVW |
| | | Non-ischemic CMP | 1.26 (0.94; 1.68) | $1.2\times10^{-1}$ | <0.001 | 49 | MR Egger |
| | | AF | 0.99 (0.97; 1.01) | $2.5\times10^{-1}$ | <0.001 | 50 | IVW |
| LV – EDM | 20 gram | Heart failure | 0.94 (0.83; 1.07) | $3.7\times10^{-1}$ | <0.001 | 20 | IVW |
| | | DCM | 1.75 (0.96; 3.19) | $6.8\times10^{-2}$ | 0.002 | 21 | IVW |
| | | Non-ischemic CMP | 0.79 (0.44; 1.44) | $4.5\times10^{-1}$ | 0.105 | 20 | IVW |
| | | AF | 0.60 (0.52; 0.68) | $6.3\times10^{-14}$ | <0.001 | 19 | IVW |
| LV – MVR | 0.1 gram/ml | Heart failure | 0.83 (0.79; 0.88) | $3.2\times10^{-11}$ | 0.006 | 59 | IVW |
| | | DCM | 0.26 (0.20; 0.34) | $1.0\times10^{-100}$ | <0.001 | 57 | IVW |

**Supplementary Table 1:** Mendelian randomization results on the effects of CMR measured changes in cardiac function and structure have on incident cardiac diagnoses. *(continued)*

| CMR measure | Units | Cardiac diagnosis | OR (95%CI) | P-value | Q p-value | No. variants | MR model |
|---|---|---|---|---|---|---|---|
| | | Non-ischemic CMP | 0.44 (0.35; 0.57) | $1.4\times10^{-10}$ | 0.242 | 56 | IVW |
| | | AF | 0.97 (0.92; 1.04) | $4.1\times10^{-1}$ | <0.001 | 46 | IVW |
| LV – TM | SD | Heart failure | 1.56 (1.02; 2.37) | $3.9\times10^{-2}$ | <0.001 | 536 | IVW |
| | | DCM | | | | | |
| | | Non-ischemic CMP | 5.07 (0.68; 37.58) | $1.1\times10^{-1}$ | 0.097 | 481 | IVW |
| | | AF | 1.10 (0.43; 2.86) | $8.4\times10^{-1}$ | <0.001 | 557 | MR Egger |
| LA – V (max) | SD | Heart failure | 1.10 (0.99; 1.21) | $8.3\times10^{-2}$ | 0.167 | 18 | IVW |
| | | DCM | 1.00 (0.63; 1.60) | $9.9\times10^{-1}$ | 0.161 | 21 | IVW |
| | | Non-ischemic CMP | 1.53 (1.00; 2.33) | $5.0\times10^{-2}$ | 0.149 | 20 | IVW |
| | | AF | 0.99 (0.91; 1.06) | $7.1\times10^{-1}$ | <0.001 | 20 | IVW |
| LA – V (min) | SD | Heart failure | 1.12 (1.00; 1.26) | $4.6\times10^{-2}$ | 0.539 | 11 | IVW |
| | | DCM | 1.69 (0.81; 3.52) | $1.7\times10^{-1}$ | 0.226 | 11 | IVW |
| | | Non-ischemic CMP | 1.31 (0.66; 2.58) | $4.4\times10^{-1}$ | 0.133 | 11 | IVW |
| | | AF | 1.18 (1.00; 1.40) | $4.5\times10^{-2}$ | 0.215 | 9 | IVW |
| LA – TF | SD | Heart failure | 0.62 (0.34; 1.12) | $1.1\times10^{-1}$ | 0.419 | 12 | MR Egger |
| | | DCM | | | | | |
| | | Non-ischemic CMP | 0.51 (0.01; 18.88) | $7.2\times10^{-1}$ | 0.366 | 10 | MR Egger |
| | | AF | 0.83 (0.72; 0.97) | $1.8\times10^{-2}$ | 0.340 | 9 | IVW |
| LA – AF | SD | Heart failure | 0.85 (0.78; 0.92) | $1.7\times10^{-4}$ | 0.624 | 21 | IVW |
| | | DCM | 0.73 (0.43; 1.24) | $2.5\times10^{-1}$ | 0.029 | 20 | IVW |
| | | Non-ischemic CMP | 1.08 (0.66; 1.75) | $7.6\times10^{-1}$ | 0.195 | 20 | IVW |
| | | AF | 0.77 (0.70; 0.85) | $2.1\times10^{-7}$ | 0.014 | 15 | IVW |
| LA – PF | SD | Heart failure | 1.05 (0.81; 1.35) | $7.1\times10^{-1}$ | 0.106 | 5 | IVW |
| | | DCM | 0.09 (0.00; 1.53) | $9.4\times10^{-2}$ | 0.650 | 6 | MR Egger |
| | | Non-ischemic CMP | 8.10 (0.70; 93.39) | $9.4\times10^{-2}$ | 0.255 | 7 | MR Egger |
| | | AF | 2.14 (1.01; 4.53) | $4.7\times10^{-2}$ | 0.075 | 5 | MR Egger |
| LA – PFR | 20 ml/s | Heart failure | 0.99 (0.91; 1.08) | $8.7\times10^{-1}$ | 0.674 | 2 | IVW |
| | | DCM | 1.44 (0.79; 2.64) | $2.4\times10^{-1}$ | None | 1 | Wald |
| | | Non-ischemic CMP | 0.80 (0.67; 0.95) | $9.9\times10^{-3}$ | 0.776 | 2 | IVW |
| | | AF | 1.01 (0.97; 1.07) | $5.7\times10^{-1}$ | 0.359 | 2 | IVW |
| RA – PFR | 20 ml/s | Heart failure | 1.01 (0.97; 1.06) | $5.8\times10^{-1}$ | 0.162 | 5 | IVW |
| | | DCM | 1.01 (0.75; 1.35) | $9.6\times10^{-1}$ | 0.065 | 5 | IVW |
| | | Non-ischemic CMP | 1.01 (0.87; 1.18) | $8.7\times10^{-1}$ | 0.022 | 5 | IVW |
| | | AF | 0.96 (0.87; 1.05) | $3.5\times10^{-1}$ | None | 1 | Wald |

*General:*
CMR: Cardiac MRI, AF: atrial fibrillation, CMP: cardiomyopathy, DCM: dilated cardiomyopathy, MR: Mendelian randomization, OR: odds ratio difference, CI: confidence interval, Q: Q test for heterogeneity. Effect estimates are coded towards the CMR increasing direction.

**Supplementary Table 2:** Mendelian randomization phewas results of the effects of CMR measured changes in funtion and structure have on disease traits.

| CMR measure | Units | Cardiac diagnosis | OR (95%CI) | P-value | Q p-value | No. variants | MR model |
|---|---|---|---|---|---|---|---|
| LV – EF | 5 % | Any stroke | 1.00 (0.94; 1.07) | $9.0\times10^{-1}$ | 0.086 | 44 | IVW |
| | | Any ischemic stroke | 1.00 (0.94; 1.06) | $9.8\times10^{-1}$ | 0.023 | 45 | IVW |
| | | Large artery stroke | 0.89 (0.76; 1.04) | $1.5\times10^{-1}$ | 0.210 | 45 | IVW |
| | | Cardioembolic stroke | 1.33 (0.72; 2.46) | $3.6\times10^{-1}$ | 0.178 | 43 | MR Egger |
| | | Small vessel stroke | 1.05 (0.92; 1.21) | $4.6\times10^{-1}$ | 0.692 | 46 | IVW |
| | | VTE | 0.96 (0.90; 1.03) | $2.2\times10^{-1}$ | 0.216 | 41 | IVW |
| | | AAA | 0.97 (0.87; 1.08) | $5.7\times10^{-1}$ | <0.001 | 47 | IVW |
| | | SBP | -0.93 (-1.14; -0.71) | $1.0\times10^{-100}$ | <0.001 | 32 | IVW |
| | | DBP | -0.60 (-0.75; -0.45) | $1.3\times10^{-14}$ | <0.001 | 24 | IVW |
| | | BMI | 0.04 (0.03; 0.05) | $5.1\times10^{-11}$ | 0.014 | 38 | IVW |
| | | T2DM | 1.00 (0.96; 1.04) | $9.7\times10^{-1}$ | 0.002 | 35 | IVW |
| | | Glycated haemoglobin | -0.01 (-0.39; 0.37) | $9.7\times10^{-1}$ | <0.001 | 40 | MR Egger |
| | | CRP | 0.01 (-0.02; 0.03) | $6.6\times10^{-1}$ | <0.001 | 37 | IVW |
| | | FEV1 | -0.07 (-0.12; -0.03) | $5.5\times10^{-4}$ | <0.001 | 40 | IVW |
| | | FVC | -0.09 (-0.13; -0.05) | $2.2\times10^{-5}$ | <0.001 | 41 | IVW |
| | | CKD | 0.98 (0.92; 1.04) | $5.1\times10^{-1}$ | <0.001 | 40 | IVW |
| | | eGFR | -0.00 (-0.00; 0.00) | $2.3\times10^{-1}$ | <0.001 | 35 | IVW |
| | | Alzheimer | 1.00 (0.99; 1.02) | $5.6\times10^{-1}$ | 0.387 | 42 | IVW |
| | | Alzheimer, late onset | 1.00 (0.91; 1.09) | $9.5\times10^{-1}$ | 0.034 | 45 | IVW |
| | | Lewy body dementia | 1.02 (0.82; 1.28) | $8.3\times10^{-1}$ | 0.435 | 41 | IVW |
| RV – EF | 5 % | Any stroke | 1.04 (0.96; 1.13) | $3.3\times10^{-1}$ | 0.096 | 41 | IVW |
| | | Any ischemic stroke | 1.04 (0.96; 1.13) | $3.0\times10^{-1}$ | 0.363 | 40 | IVW |
| | | Large artery stroke | 1.14 (0.92; 1.42) | $2.2\times10^{-1}$ | 0.066 | 41 | IVW |
| | | Cardioembolic stroke | 1.14 (0.97; 1.35) | $1.1\times10^{-1}$ | 0.095 | 41 | IVW |
| | | Small vessel stroke | 1.04 (0.88; 1.24) | $6.3\times10^{-1}$ | 0.589 | 41 | IVW |
| | | VTE | 0.92 (0.85; 1.00) | $4.5\times10^{-2}$ | 0.040 | 37 | IVW |
| | | AAA | 1.08 (0.95; 1.22) | $2.3\times10^{-1}$ | <0.001 | 42 | IVW |
| | | SBP | -0.10 (-0.41; 0.21) | $5.4\times10^{-1}$ | <0.001 | 21 | IVW |
| | | DBP | -0.32 (-0.50; -0.15) | $3.2\times10^{-4}$ | <0.001 | 21 | IVW |
| | | BMI | -0.09 (-0.14; -0.04) | $3.4\times10^{-4}$ | <0.001 | 28 | MR Egger |
| | | T2DM | 0.97 (0.92; 1.02) | $2.1\times10^{-1}$ | <0.001 | 29 | IVW |
| | | Glycated haemoglobin | 0.18 (0.07; 0.30) | $2.1\times10^{-3}$ | 0.002 | 42 | IVW |
| | | CRP | -0.05 (-0.16; 0.06) | $3.4\times10^{-1}$ | 0.010 | 34 | MR Egger |
| | | FEV1 | -0.04 (-0.09; 0.01) | $1.4\times10^{-1}$ | 0.003 | 41 | IVW |
| | | FVC | -0.04 (-0.09; 0.01) | $1.3\times10^{-1}$ | <0.001 | 41 | IVW |
| | | CKD | 0.92 (0.86; 0.98) | $1.1\times10^{-2}$ | <0.001 | 41 | IVW |
| | | eGFR | 0.00 (-0.00; 0.01) | $7.5\times10^{-2}$ | 0.004 | 34 | IVW |
| | | Alzheimer | 1.00 (0.98; 1.02) | $9.8\times10^{-1}$ | 0.079 | 43 | IVW |
| | | Alzheimer, late onset | 1.07 (0.96; 1.20) | $2.3\times10^{-1}$ | 0.820 | 42 | IVW |
| | | Lewy body dementia | 1.26 (0.95; 1.65) | $1.0\times10^{-1}$ | 0.352 | 37 | IVW |
| LV – SV | 20 ml | Any stroke | 0.70 (0.06; 7.50) | $7.6\times10^{-1}$ | 0.468 | 6 | MR Egger |
| | | Any ischemic stroke | 0.61 (0.06; 6.60) | $6.8\times10^{-1}$ | 0.375 | 7 | MR Egger |
| | | Large artery stroke | 2.16 (1.08; 4.31) | $2.9\times10^{-2}$ | 0.399 | 9 | IVW |
| | | Cardioembolic stroke | 0.86 (0.00; 808.61) | $9.7\times10^{-1}$ | 0.057 | 6 | MR Egger |
| | | Small vessel stroke | 0.41 (0.20; 0.85) | $1.6\times10^{-2}$ | 0.115 | 10 | IVW |
| | | VTE | 0.85 (0.67; 1.07) | $1.6\times10^{-1}$ | 0.582 | 7 | IVW |
| | | AAA | 1.33 (0.90; 1.98) | $1.5\times10^{-1}$ | 0.345 | 9 | IVW |
| | | SBP | -5.18 (-13.71; 3.35) | $2.3\times10^{-1}$ | 0.367 | 3 | MR Egger |
| | | DBP | -2.33 (-2.92; -1.73) | $2.1\times10^{-14}$ | 0.583 | 5 | IVW |
| | | BMI | -0.03 (-0.10; 0.04) | $4.5\times10^{-1}$ | 0.063 | 7 | IVW |
| | | T2DM | 0.79 (0.66; 0.94) | $8.9\times10^{-3}$ | <0.001 | 6 | IVW |
| | | Glycated haemoglobin | 0.65 (-2.59; 3.89) | $6.9\times10^{-1}$ | 0.010 | 7 | MR Egger |
| | | CRP | -0.06 (-0.16; 0.05) | $3.0\times10^{-1}$ | 0.675 | 8 | IVW |
| | | FEV1 | 1.12 (-0.74; 2.98) | $2.4\times10^{-1}$ | 0.165 | 6 | MR Egger |
| | | FVC | 0.97 (-0.71; 2.64) | $2.6\times10^{-1}$ | 0.245 | 7 | MR Egger |
| | | CKD | 0.71 (0.05; 9.67) | $8.0\times10^{-1}$ | 0.344 | 7 | MR Egger |
| | | eGFR | 0.01 (-0.00; 0.02) | $2.0\times10^{-1}$ | 0.602 | 8 | IVW |
| | | Alzheimer | 1.01 (0.63; 1.61) | $9.7\times10^{-1}$ | 0.007 | 8 | MR Egger |
| | | Alzheimer, late onset | 1.71 (0.02; 135.56) | $8.1\times10^{-1}$ | 0.061 | 9 | MR Egger |
| | | Lewy body dementia | 1.42 (0.55; 3.68) | $4.7\times10^{-1}$ | 0.878 | 7 | IVW |
| RV – SV | 20 ml | Any stroke | 0.68 (0.54; 0.85) | $9.1\times10^{-4}$ | <0.001 | 7 | IVW |
| | | Any ischemic stroke | 0.79 (0.54; 1.16) | $2.3\times10^{-1}$ | 0.068 | 6 | IVW |
| | | Large artery stroke | 0.48 (0.27; 0.85) | $1.2\times10^{-2}$ | 0.008 | 9 | IVW |
| | | Cardioembolic stroke | 0.68 (0.47; 0.99) | $4.4\times10^{-2}$ | 0.002 | 10 | IVW |
| | | Small vessel stroke | 0.43 (0.21; 0.86) | $1.7\times10^{-2}$ | 0.092 | 9 | IVW |
| | | VTE | 1.67 (0.80; 3.50) | $1.7\times10^{-1}$ | 0.006 | 12 | MR Egger |

**Supplementary Table 2:** Mendelian randomization phewas results of the effects of CMR measured changes in funtion and structure have on disease traits. *(continued)*

| CMR measure | Units | Cardiac diagnosis | OR (95%CI) | P-value | Q p-value | No. variants | MR model |
|---|---|---|---|---|---|---|---|
| | | AAA | 0.73 (0.53; 1.01) | $5.4\times10^{-2}$ | 0.025 | 12 | IVW |
| | | SBP | -6.50 (-8.47; -4.52) | $1.2\times10^{-10}$ | None | 1 | Wald |
| | | DBP | -6.40 (-7.56; -5.25) | $1.0\times10^{-100}$ | None | 1 | Wald |
| | | BMI | -0.12 (-0.27; 0.03) | $1.1\times10^{-1}$ | 0.007 | 7 | MR Egger |
| | | T2DM | 0.75 (0.65; 0.85) | $1.4\times10^{-5}$ | 0.316 | 10 | IVW |
| | | Glycated haemoglobin | -2.04 (-3.57; -0.51) | $8.9\times10^{-3}$ | 0.044 | 9 | MR Egger |
| | | CRP | -0.07 (-0.15; 0.01) | $6.8\times10^{-2}$ | 0.838 | 11 | IVW |
| | | FEV1 | 0.07 (-0.07; 0.20) | $3.2\times10^{-1}$ | 0.574 | 12 | IVW |
| | | FVC | 0.05 (-0.09; 0.19) | $5.1\times10^{-1}$ | 0.331 | 12 | IVW |
| | | CKD | 3.22 (0.89; 11.68) | $7.6\times10^{-2}$ | 0.053 | 10 | MR Egger |
| | | eGFR | -0.03 (-0.07; 0.01) | $1.0\times10^{-1}$ | <0.001 | 9 | MR Egger |
| | | Alzheimer | 1.02 (0.97; 1.08) | $4.0\times10^{-1}$ | 0.417 | 11 | IVW |
| | | Alzheimer, late onset | 0.32 (0.11; 0.98) | $4.6\times10^{-2}$ | 0.025 | 11 | MR Egger |
| | | Lewy body dementia | 3.05 (1.46; 6.37) | $3.0\times10^{-3}$ | 0.014 | 9 | IVW |
| LV – PFR | 10 ml/s | Any stroke | 0.96 (0.92; 1.00) | $7.3\times10^{-2}$ | 0.371 | 2 | IVW |
| | | Any ischemic stroke | 0.95 (0.91; 1.00) | $5.3\times10^{-2}$ | 0.439 | 2 | IVW |
| | | Large artery stroke | 0.93 (0.82; 1.05) | $2.2\times10^{-1}$ | 0.534 | 2 | IVW |
| | | Cardioembolic stroke | 0.97 (0.88; 1.07) | $5.2\times10^{-1}$ | 0.841 | 2 | IVW |
| | | Small vessel stroke | 1.00 (0.89; 1.12) | $1.0\times10^{0}$ | 0.601 | 2 | IVW |
| | | VTE | 0.99 (0.96; 1.03) | $7.7\times10^{-1}$ | 0.584 | 2 | IVW |
| | | AAA | 0.99 (0.93; 1.06) | $8.3\times10^{-1}$ | 0.321 | 2 | IVW |
| | | SBP | 0.09 (-0.08; 0.26) | $3.1\times10^{-1}$ | None | 1 | Wald |
| | | DBP | 0.02 (-0.08; 0.12) | $6.4\times10^{-1}$ | None | 1 | Wald |
| | | BMI | 0.00 (-0.00; 0.01) | $4.0\times10^{-1}$ | 0.735 | 2 | IVW |
| | | T2DM | | | | | |
| | | Glycated haemoglobin | | | | | |
| | | CRP | -0.01 (-0.03; 0.02) | $5.5\times10^{-1}$ | None | 1 | Wald |
| | | FEV1 | 0.00 (-0.02; 0.03) | $8.0\times10^{-1}$ | 0.740 | 2 | IVW |
| | | FVC | 0.00 (-0.03; 0.03) | $9.1\times10^{-1}$ | 0.260 | 2 | IVW |
| | | CKD | 1.02 (0.98; 1.07) | $3.9\times10^{-1}$ | 0.975 | 2 | IVW |
| | | eGFR | -0.00 (-0.00; 0.00) | $9.9\times10^{-1}$ | 0.240 | 2 | IVW |
| | | Alzheimer | 1.00 (0.99; 1.02) | $5.8\times10^{-1}$ | 0.310 | 2 | IVW |
| | | Alzheimer, late onset | 1.20 (1.10; 1.31) | $7.0\times10^{-5}$ | None | 1 | Wald |
| | | Lewy body dementia | 0.97 (0.84; 1.11) | $6.6\times10^{-1}$ | 0.757 | 2 | IVW |
| RV – PFR | 10 ml/s | Any stroke | 0.96 (0.91; 1.00) | $6.0\times10^{-2}$ | 0.531 | 2 | IVW |
| | | Any ischemic stroke | 0.95 (0.90; 1.00) | $6.4\times10^{-2}$ | 0.511 | 2 | IVW |
| | | Large artery stroke | 0.89 (0.75; 1.06) | $2.0\times10^{-1}$ | 0.219 | 2 | IVW |
| | | Cardioembolic stroke | 1.03 (0.92; 1.15) | $5.8\times10^{-1}$ | 0.759 | 2 | IVW |
| | | Small vessel stroke | 0.87 (0.77; 0.99) | $2.9\times10^{-2}$ | 0.977 | 2 | IVW |
| | | VTE | 1.04 (1.00; 1.08) | $5.0\times10^{-2}$ | 0.939 | 2 | IVW |
| | | AAA | 0.93 (0.86; 1.01) | $8.5\times10^{-2}$ | 0.881 | 2 | IVW |
| | | SBP | -6.26 (-8.92; -3.61) | $3.9\times10^{-6}$ | None | 2 | MR Egger |
| | | DBP | -3.18 (-4.71; -1.66) | $4.1\times10^{-5}$ | None | 2 | MR Egger |
| | | BMI | -0.00 (-0.01; 0.01) | $4.1\times10^{-1}$ | 0.225 | 2 | IVW |
| | | T2DM | 0.37 (0.21; 0.64) | $4.2\times10^{-4}$ | None | 2 | MR Egger |
| | | Glycated haemoglobin | -2.79 (-4.09; -1.49) | $2.5\times10^{-5}$ | None | 2 | MR Egger |
| | | CRP | -0.01 (-0.03; 0.01) | $3.5\times10^{-1}$ | 0.733 | 2 | IVW |
| | | FEV1 | -0.02 (-0.07; 0.02) | $3.0\times10^{-1}$ | 0.123 | 2 | IVW |
| | | FVC | -0.02 (-0.05; 0.01) | $1.9\times10^{-1}$ | 0.268 | 2 | IVW |
| | | CKD | 0.95 (0.91; 1.00) | $6.6\times10^{-2}$ | 0.243 | 2 | IVW |
| | | eGFR | 0.00 (0.00; 0.00) | $1.5\times10^{-2}$ | 0.728 | 2 | IVW |
| | | Alzheimer | 1.00 (1.00; 1.01) | $3.2\times10^{-1}$ | 0.613 | 2 | IVW |
| | | Alzheimer, late onset | 0.94 (0.88; 1.00) | $6.7\times10^{-2}$ | 0.467 | 2 | IVW |
| | | Lewy body dementia | 0.95 (0.77; 1.18) | $6.5\times10^{-1}$ | None | 1 | Wald |
| LV – PER | 20 ml/s | Any stroke | 1.00 (0.93; 1.06) | $8.8\times10^{-1}$ | 0.502 | 3 | IVW |
| | | Any ischemic stroke | 1.02 (0.95; 1.09) | $5.8\times10^{-1}$ | 0.416 | 3 | IVW |
| | | Large artery stroke | 1.06 (0.89; 1.25) | $5.3\times10^{-1}$ | 0.466 | 3 | IVW |
| | | Cardioembolic stroke | 1.14 (0.96; 1.36) | $1.3\times10^{-1}$ | 0.160 | 3 | IVW |
| | | Small vessel stroke | 1.01 (0.84; 1.22) | $9.1\times10^{-1}$ | 0.240 | 3 | IVW |
| | | VTE | 1.02 (0.97; 1.07) | $4.1\times10^{-1}$ | 0.552 | 4 | IVW |
| | | AAA | 0.95 (0.88; 1.03) | $1.9\times10^{-1}$ | 0.950 | 3 | IVW |
| | | SBP | 0.20 (-0.04; 0.45) | $1.0\times10^{-1}$ | 0.344 | 2 | IVW |
| | | DBP | 0.08 (-0.11; 0.28) | $4.0\times10^{-1}$ | None | 1 | Wald |
| | | BMI | -0.04 (-0.06; -0.02) | $8.6\times10^{-5}$ | None | 1 | Wald |
| | | T2DM | 0.94 (0.90; 0.98) | $6.9\times10^{-3}$ | 0.334 | 3 | IVW |
| | | Glycated haemoglobin | -0.04 (-0.12; 0.04) | $3.5\times10^{-1}$ | 0.320 | 4 | IVW |

**Supplementary Table 2:** Mendelian randomization phewas results of the effects of CMR measured changes in funtion and structure have on disease traits. *(continued)*

| CMR measure | Units | Cardiac diagnosis | OR (95%CI) | P-value | Q p-value | No. variants | MR model |
|---|---|---|---|---|---|---|---|
| | | CRP | -0.02 (-0.06; 0.02) | $3.8\times10^{-1}$ | 0.082 | 3 | IVW |
| | | FEV1 | -0.01 (-0.04; 0.02) | $6.6\times10^{-1}$ | 0.574 | 4 | IVW |
| | | FVC | 0.00 (-0.03; 0.03) | $9.1\times10^{-1}$ | 0.615 | 4 | IVW |
| | | CKD | 1.03 (0.97; 1.10) | $3.4\times10^{-1}$ | 0.669 | 3 | IVW |
| | | eGFR | -0.00 (-0.01; -0.00) | $2.5\times10^{-2}$ | 0.367 | 3 | IVW |
| | | Alzheimer | 1.00 (0.97; 1.02) | $7.7\times10^{-1}$ | 0.097 | 2 | IVW |
| | | Alzheimer, late onset | 0.99 (0.92; 1.06) | $7.2\times10^{-1}$ | 0.049 | 3 | IVW |
| | | Lewy body dementia | 1.15 (0.97; 1.37) | $1.2\times10^{-1}$ | 0.376 | 4 | IVW |
| RV – PER | 20 ml/s | Any stroke | 1.02 (0.97; 1.08) | $4.3\times10^{-1}$ | 0.274 | 7 | IVW |
| | | Any ischemic stroke | 1.04 (0.99; 1.10) | $1.5\times10^{-1}$ | 0.290 | 7 | IVW |
| | | Large artery stroke | 0.94 (0.84; 1.06) | $3.2\times10^{-1}$ | <0.001 | 8 | IVW |
| | | Cardioembolic stroke | 0.94 (0.84; 1.06) | $3.0\times10^{-1}$ | 0.102 | 8 | IVW |
| | | Small vessel stroke | 1.01 (0.90; 1.13) | $8.7\times10^{-1}$ | 0.454 | 7 | IVW |
| | | VTE | 0.97 (0.93; 1.01) | $1.3\times10^{-1}$ | 0.283 | 8 | IVW |
| | | AAA | 1.05 (0.98; 1.13) | $1.3\times10^{-1}$ | 0.003 | 9 | IVW |
| | | SBP | -0.25 (-0.48; -0.02) | $3.1\times10^{-2}$ | 0.323 | 3 | IVW |
| | | DBP | -0.30 (-0.55; -0.05) | $1.8\times10^{-2}$ | 0.105 | 2 | IVW |
| | | BMI | -0.00 (-0.01; 0.01) | $8.1\times10^{-1}$ | 0.011 | 5 | IVW |
| | | T2DM | 1.02 (0.99; 1.04) | $2.4\times10^{-1}$ | 0.020 | 7 | IVW |
| | | Glycated haemoglobin | -0.34 (-0.54; -0.14) | $6.9\times10^{-4}$ | 0.002 | 7 | MR Egger |
| | | CRP | -0.01 (-0.03; 0.02) | $5.7\times10^{-1}$ | 0.408 | 5 | IVW |
| | | FEV1 | -0.03 (-0.05; -0.00) | $2.8\times10^{-2}$ | 0.010 | 8 | IVW |
| | | FVC | -0.01 (-0.04; 0.02) | $4.7\times10^{-1}$ | 0.806 | 7 | IVW |
| | | CKD | 0.93 (0.89; 0.96) | $9.7\times10^{-5}$ | 0.046 | 8 | IVW |
| | | eGFR | -0.00 (-0.01; 0.00) | $4.6\times10^{-1}$ | 0.002 | 8 | MR Egger |
| | | Alzheimer | 0.96 (0.94; 0.99) | $1.3\times10^{-2}$ | 0.670 | 9 | MR Egger |
| | | Alzheimer, late onset | 1.02 (0.95; 1.09) | $6.5\times10^{-1}$ | 0.217 | 8 | IVW |
| | | Lewy body dementia | 1.04 (0.89; 1.23) | $6.1\times10^{-1}$ | 0.654 | 8 | IVW |
| LV – EDV | 20 ml | Any stroke | 0.95 (0.90; 1.01) | $1.1\times10^{-1}$ | 0.027 | 28 | IVW |
| | | Any ischemic stroke | 0.99 (0.93; 1.06) | $7.5\times10^{-1}$ | 0.026 | 28 | IVW |
| | | Large artery stroke | 1.04 (0.87; 1.25) | $6.5\times10^{-1}$ | 0.125 | 29 | IVW |
| | | Cardioembolic stroke | 0.81 (0.72; 0.92) | $9.8\times10^{-4}$ | <0.001 | 29 | IVW |
| | | Small vessel stroke | 0.98 (0.85; 1.14) | $8.2\times10^{-1}$ | 0.947 | 29 | IVW |
| | | VTE | 1.04 (0.97; 1.11) | $2.9\times10^{-1}$ | 0.414 | 27 | IVW |
| | | AAA | 0.93 (0.81; 1.07) | $3.2\times10^{-1}$ | 0.207 | 29 | IVW |
| | | SBP | 0.77 (0.54; 0.99) | $3.1\times10^{-11}$ | 0.024 | 22 | IVW |
| | | DBP | -0.32 (-0.50; -0.14) | $5.1\times10^{-4}$ | 0.069 | 17 | IVW |
| | | BMI | 0.00 (-0.06; 0.06) | $9.6\times10^{-1}$ | 0.002 | 24 | MR Egger |
| | | T2DM | 0.95 (0.90; 0.99) | $1.9\times10^{-2}$ | <0.001 | 25 | IVW |
| | | Glycated haemoglobin | -0.15 (-0.27; -0.03) | $1.3\times10^{-2}$ | 0.036 | 26 | IVW |
| | | CRP | 0.01 (-0.02; 0.03) | $6.4\times10^{-1}$ | 0.002 | 29 | IVW |
| | | FEV1 | 0.05 (0.01; 0.09) | $1.3\times10^{-2}$ | <0.001 | 32 | IVW |
| | | FVC | 0.08 (0.04; 0.12) | $7.2\times10^{-5}$ | 0.008 | 31 | IVW |
| | | CKD | 0.95 (0.89; 1.01) | $1.0\times10^{-1}$ | 0.007 | 28 | IVW |
| | | eGFR | -0.00 (-0.00; 0.00) | $6.9\times10^{-2}$ | 0.011 | 24 | IVW |
| | | Alzheimer | 1.02 (1.00; 1.04) | $1.1\times10^{-1}$ | 0.073 | 27 | IVW |
| | | Alzheimer, late onset | 0.82 (0.75; 0.90) | $2.3\times10^{-5}$ | <0.001 | 29 | IVW |
| | | Lewy body dementia | 11.86 (2.08; 67.63) | $5.3\times10^{-3}$ | 0.167 | 25 | MR Egger |
| RV – EDV | 20 ml | Any stroke | 1.08 (1.01; 1.15) | $2.8\times10^{-2}$ | <0.001 | 32 | IVW |
| | | Any ischemic stroke | 1.07 (1.00; 1.16) | $5.5\times10^{-2}$ | 0.005 | 30 | IVW |
| | | Large artery stroke | 0.98 (0.83; 1.16) | $8.3\times10^{-1}$ | <0.001 | 36 | IVW |
| | | Cardioembolic stroke | 0.84 (0.75; 0.95) | $3.5\times10^{-3}$ | <0.001 | 37 | IVW |
| | | Small vessel stroke | 0.86 (0.74; 1.00) | $4.7\times10^{-2}$ | 0.007 | 37 | IVW |
| | | VTE | 0.86 (0.81; 0.91) | $1.5\times10^{-6}$ | <0.001 | 37 | IVW |
| | | AAA | 0.87 (0.78; 0.97) | $1.6\times10^{-2}$ | 0.127 | 43 | IVW |
| | | SBP | -0.47 (-0.80; -0.13) | $6.8\times10^{-3}$ | 0.004 | 12 | IVW |
| | | DBP | -1.53 (-1.76; -1.31) | $1.0\times10^{-100}$ | <0.001 | 10 | IVW |
| | | BMI | 0.03 (0.02; 0.04) | $5.5\times10^{-6}$ | <0.001 | 32 | IVW |
| | | T2DM | 0.88 (0.84; 0.91) | $4.2\times10^{-10}$ | <0.001 | 32 | IVW |
| | | Glycated haemoglobin | 0.11 (0.00; 0.22) | $4.5\times10^{-2}$ | <0.001 | 35 | IVW |
| | | CRP | -0.04 (-0.07; -0.02) | $1.7\times10^{-3}$ | 0.008 | 38 | IVW |
| | | FEV1 | 0.04 (0.00; 0.08) | $3.6\times10^{-2}$ | 0.332 | 42 | IVW |
| | | FVC | 0.01 (-0.03; 0.05) | $5.2\times10^{-1}$ | 0.009 | 42 | IVW |
| | | CKD | 0.99 (0.92; 1.05) | $6.6\times10^{-1}$ | <0.001 | 33 | IVW |
| | | eGFR | -0.00 (-0.00; 0.00) | $9.3\times10^{-1}$ | <0.001 | 30 | IVW |
| | | Alzheimer | 1.01 (0.99; 1.02) | $3.2\times10^{-1}$ | 0.047 | 44 | IVW |

**Supplementary Table 2:** Mendelian randomization phewas results of the effects of CMR measured changes in funtion and structure have on disease traits. *(continued)*

| CMR measure | Units | Cardiac diagnosis | OR (95%CI) | P-value | Q p-value | No. variants | MR model |
|---|---|---|---|---|---|---|---|
| | | Alzheimer, late onset | 1.04 (0.96; 1.13) | $3.1\times10^{-1}$ | <0.001 | 41 | IVW |
| | | Lewy body dementia | 1.34 (1.07; 1.67) | $1.0\times10^{-2}$ | 0.004 | 38 | IVW |
| LV – ESV | 5 ml | Any stroke | 0.99 (0.97; 1.01) | $3.8\times10^{-1}$ | 0.102 | 56 | IVW |
| | | Any ischemic stroke | 1.00 (0.98; 1.03) | $7.7\times10^{-1}$ | 0.023 | 56 | IVW |
| | | Large artery stroke | 1.06 (1.00; 1.12) | $6.9\times10^{-2}$ | 0.013 | 55 | IVW |
| | | Cardioembolic stroke | 0.93 (0.89; 0.97) | $4.7\times10^{-4}$ | 0.002 | 57 | IVW |
| | | Small vessel stroke | 1.00 (0.95; 1.06) | $8.9\times10^{-1}$ | 0.952 | 56 | IVW |
| | | VTE | 1.02 (0.99; 1.04) | $1.9\times10^{-1}$ | 0.051 | 56 | IVW |
| | | AAA | 1.06 (1.02; 1.10) | $2.9\times10^{-3}$ | 0.002 | 62 | IVW |
| | | SBP | 0.24 (0.16; 0.32) | $1.6\times10^{-8}$ | <0.001 | 42 | IVW |
| | | DBP | 0.07 (0.02; 0.12) | $4.2\times10^{-3}$ | <0.001 | 41 | IVW |
| | | BMI | 0.00 (-0.00; 0.01) | $1.3\times10^{-1}$ | <0.001 | 52 | IVW |
| | | T2DM | 0.99 (0.98; 1.01) | $4.0\times10^{-1}$ | <0.001 | 52 | IVW |
| | | Glycated haemoglobin | -0.04 (-0.08; -0.01) | $1.4\times10^{-2}$ | <0.001 | 59 | IVW |
| | | CRP | -0.01 (-0.02; 0.00) | $2.3\times10^{-1}$ | 0.014 | 58 | IVW |
| | | FEV1 | 0.04 (0.02; 0.05) | $5.5\times10^{-8}$ | 0.014 | 59 | IVW |
| | | FVC | 0.03 (0.02; 0.05) | $1.5\times10^{-6}$ | 0.014 | 60 | IVW |
| | | CKD | 0.99 (0.97; 1.01) | $4.9\times10^{-1}$ | 0.043 | 55 | IVW |
| | | eGFR | -0.00 (-0.00; 0.00) | $8.1\times10^{-1}$ | <0.001 | 49 | IVW |
| | | Alzheimer | 1.00 (1.00; 1.01) | $5.5\times10^{-1}$ | 0.023 | 56 | IVW |
| | | Alzheimer, late onset | 0.95 (0.92; 0.98) | $2.7\times10^{-3}$ | <0.001 | 57 | IVW |
| | | Lewy body dementia | 1.05 (0.96; 1.15) | $2.9\times10^{-1}$ | 0.324 | 54 | IVW |
| RV – ESV | 5 ml | Any stroke | 1.02 (1.00; 1.04) | $6.1\times10^{-2}$ | <0.001 | 51 | IVW |
| | | Any ischemic stroke | 0.89 (0.78; 1.01) | $7.0\times10^{-2}$ | 0.019 | 38 | MR Egger |
| | | Large artery stroke | 1.03 (0.77; 1.37) | $8.4\times10^{-1}$ | 0.026 | 50 | MR Egger |
| | | Cardioembolic stroke | 0.99 (0.95; 1.03) | $6.5\times10^{-1}$ | 0.006 | 52 | IVW |
| | | Small vessel stroke | 1.00 (0.95; 1.05) | $9.9\times10^{-1}$ | 0.405 | 52 | IVW |
| | | VTE | 1.01 (0.99; 1.04) | $1.7\times10^{-1}$ | 0.002 | 51 | IVW |
| | | AAA | 0.99 (0.95; 1.03) | $7.0\times10^{-1}$ | 0.245 | 55 | IVW |
| | | SBP | 0.30 (0.22; 0.38) | $6.4\times10^{-13}$ | <0.001 | 33 | IVW |
| | | DBP | 0.03 (-0.03; 0.08) | $3.3\times10^{-1}$ | <0.001 | 27 | IVW |
| | | BMI | 0.01 (0.00; 0.01) | $5.0\times10^{-5}$ | <0.001 | 43 | IVW |
| | | T2DM | 1.02 (1.01; 1.04) | $3.9\times10^{-3}$ | <0.001 | 46 | IVW |
| | | Glycated haemoglobin | -0.01 (-0.05; 0.02) | $5.6\times10^{-1}$ | <0.001 | 51 | IVW |
| | | CRP | -0.01 (-0.02; 0.00) | $5.9\times10^{-2}$ | 0.022 | 52 | IVW |
| | | FEV1 | 0.01 (-0.00; 0.03) | $7.0\times10^{-2}$ | 0.027 | 53 | IVW |
| | | FVC | 0.01 (-0.00; 0.03) | $1.1\times10^{-1}$ | 0.022 | 53 | IVW |
| | | CKD | 1.02 (1.00; 1.04) | $1.1\times10^{-1}$ | <0.001 | 48 | IVW |
| | | eGFR | -0.00 (-0.00; -0.00) | $1.8\times10^{-2}$ | <0.001 | 45 | IVW |
| | | Alzheimer | 1.00 (1.00; 1.01) | $2.3\times10^{-1}$ | 0.011 | 56 | IVW |
| | | Alzheimer, late onset | 0.98 (0.95; 1.01) | $1.2\times10^{-1}$ | <0.001 | 54 | IVW |
| | | Lewy body dementia | 1.03 (0.95; 1.11) | $4.8\times10^{-1}$ | <0.001 | 51 | IVW |
| LV – EDM | 20 gram | Any stroke | 0.87 (0.74; 1.02) | $8.1\times10^{-2}$ | 0.048 | 16 | IVW |
| | | Any ischemic stroke | 0.87 (0.71; 1.08) | $2.1\times10^{-1}$ | 0.085 | 16 | IVW |
| | | Large artery stroke | 1.24 (0.84; 1.83) | $2.7\times10^{-1}$ | 0.518 | 19 | IVW |
| | | Cardioembolic stroke | 0.87 (0.64; 1.16) | $3.4\times10^{-1}$ | <0.001 | 20 | IVW |
| | | Small vessel stroke | 1.02 (0.71; 1.46) | $9.0\times10^{-1}$ | 0.009 | 19 | IVW |
| | | VTE | 1.08 (0.89; 1.32) | $4.2\times10^{-1}$ | 0.075 | 21 | IVW |
| | | AAA | 0.85 (0.64; 1.12) | $2.5\times10^{-1}$ | 0.006 | 23 | IVW |
| | | SBP | -0.32 (-0.83; 0.20) | $2.3\times10^{-1}$ | 0.034 | 14 | IVW |
| | | DBP | -0.57 (-0.88; -0.25) | $3.8\times10^{-4}$ | <0.001 | 13 | IVW |
| | | BMI | -0.01 (-0.04; 0.02) | $4.2\times10^{-1}$ | <0.001 | 17 | IVW |
| | | T2DM | 0.70 (0.63; 0.76) | $5.8\times10^{-14}$ | <0.001 | 20 | IVW |
| | | Glycated haemoglobin | -0.40 (-0.60; -0.20) | $9.1\times10^{-5}$ | 0.037 | 24 | IVW |
| | | CRP | 0.46 (0.14; 0.78) | $5.4\times10^{-3}$ | 0.286 | 20 | MR Egger |
| | | FEV1 | 0.10 (0.00; 0.19) | $4.2\times10^{-2}$ | 0.242 | 23 | IVW |
| | | FVC | 0.11 (0.03; 0.20) | $9.5\times10^{-3}$ | 0.609 | 23 | IVW |
| | | CKD | 0.99 (0.85; 1.16) | $9.4\times10^{-1}$ | 0.011 | 17 | IVW |
| | | eGFR | -0.01 (-0.01; -0.00) | $8.1\times10^{-3}$ | <0.001 | 15 | IVW |
| | | Alzheimer | 0.98 (0.95; 1.02) | $4.0\times10^{-1}$ | 0.286 | 22 | IVW |
| | | Alzheimer, late onset | 0.68 (0.55; 0.84) | $4.8\times10^{-4}$ | 0.030 | 21 | IVW |
| | | Lewy body dementia | 1.60 (0.78; 3.28) | $2.0\times10^{-1}$ | 0.068 | 20 | IVW |
| LV – MVR | 0.1 gram/ml | Any stroke | 0.87 (0.81; 0.93) | $2.7\times10^{-5}$ | <0.001 | 57 | IVW |
| | | Any ischemic stroke | 0.81 (0.76; 0.87) | $1.9\times10^{-8}$ | <0.001 | 58 | IVW |
| | | Large artery stroke | 0.75 (0.63; 0.89) | $1.2\times10^{-3}$ | <0.001 | 56 | IVW |
| | | Cardioembolic stroke | 0.63 (0.55; 0.73) | $6.8\times10^{-11}$ | <0.001 | 57 | IVW |

**Supplementary Table 2:** Mendelian randomization phewas results of the effects of CMR measured changes in funtion and structure have on disease traits. *(continued)*

| CMR measure | Units | Cardiac diagnosis | OR (95%CI) | P-value | Q p-value | No. variants | MR model |
|---|---|---|---|---|---|---|---|
| | | Small vessel stroke | 0.85 (0.72; 0.99) | $4.3\times10^{-2}$ | <0.001 | 57 | IVW |
| | | VTE | 1.21 (1.13; 1.30) | $4.8\times10^{-8}$ | <0.001 | 54 | IVW |
| | | AAA | 1.04 (0.91; 1.19) | $5.4\times10^{-1}$ | 0.022 | 58 | IVW |
| | | SBP | -0.97 (-1.21; -0.73) | $5.6\times10^{-15}$ | <0.001 | 44 | IVW |
| | | DBP | -0.96 (-1.11; -0.80) | $1.0\times10^{-100}$ | <0.001 | 38 | IVW |
| | | BMI | -0.00 (-0.01; 0.01) | $9.8\times10^{-1}$ | <0.001 | 52 | IVW |
| | | T2DM | 0.88 (0.84; 0.93) | $2.8\times10^{-6}$ | <0.001 | 47 | IVW |
| | | Glycated haemoglobin | -0.01 (-0.14; 0.12) | $8.8\times10^{-1}$ | <0.001 | 52 | IVW |
| | | CRP | -0.07 (-0.10; -0.04) | $2.1\times10^{-6}$ | <0.001 | 53 | IVW |
| | | FEV1 | -0.07 (-0.11; -0.03) | $2.0\times10^{-3}$ | <0.001 | 49 | IVW |
| | | FVC | -0.01 (-0.06; 0.03) | $5.9\times10^{-1}$ | <0.001 | 51 | IVW |
| | | CKD | 1.07 (1.00; 1.14) | $3.8\times10^{-2}$ | <0.001 | 57 | IVW |
| | | eGFR | -0.01 (-0.02; -0.01) | $1.0\times10^{-100}$ | <0.001 | 49 | IVW |
| | | Alzheimer | 1.00 (0.98; 1.02) | $9.7\times10^{-1}$ | 0.002 | 65 | IVW |
| | | Alzheimer, late onset | 1.09 (0.98; 1.20) | $1.2\times10^{-1}$ | 0.003 | 55 | IVW |
| | | Lewy body dementia | 0.75 (0.57; 0.98) | $3.4\times10^{-2}$ | 0.010 | 52 | IVW |
| LV – TM | SD | Any stroke | 0.36 (0.08; 1.62) | $1.8\times10^{-1}$ | 0.189 | 480 | MR Egger |
| | | Any ischemic stroke | 0.48 (0.09; 2.48) | $3.8\times10^{-1}$ | 0.239 | 479 | MR Egger |
| | | Large artery stroke | 0.38 (0.01; 20.92) | $6.4\times10^{-1}$ | <0.001 | 478 | MR Egger |
| | | Cardioembolic stroke | 0.02 (0.00; 0.47) | $1.6\times10^{-2}$ | 0.044 | 473 | MR Egger |
| | | Small vessel stroke | 1.41 (0.32; 6.30) | $6.5\times10^{-1}$ | 0.217 | 477 | IVW |
| | | VTE | 1.29 (0.74; 2.23) | $3.7\times10^{-1}$ | 0.005 | 527 | IVW |
| | | AAA | 1.03 (0.39; 2.73) | $9.5\times10^{-1}$ | 0.047 | 547 | IVW |
| | | SBP | 7.23 (1.98; 12.48) | $6.9\times10^{-3}$ | <0.001 | 368 | MR Egger |
| | | DBP | 1.44 (0.28; 2.61) | $1.5\times10^{-2}$ | <0.001 | 377 | IVW |
| | | BMI | 0.20 (-0.05; 0.45) | $1.2\times10^{-1}$ | <0.001 | 461 | MR Egger |
| | | T2DM | 1.17 (0.80; 1.70) | $4.2\times10^{-1}$ | <0.001 | 472 | IVW |
| | | Glycated haemoglobin | 1.05 (0.23; 1.86) | $1.2\times10^{-2}$ | <0.001 | 547 | IVW |
| | | CRP | 0.09 (-0.18; 0.36) | $5.3\times10^{-1}$ | 0.007 | 438 | IVW |
| | | FEV1 | -0.06 (-0.42; 0.30) | $7.4\times10^{-1}$ | 0.022 | 538 | IVW |
| | | FVC | -0.06 (-0.42; 0.29) | $7.3\times10^{-1}$ | 0.020 | 537 | IVW |
| | | CKD | 0.90 (0.50; 1.61) | $7.3\times10^{-1}$ | 0.009 | 467 | IVW |
| | | eGFR | 0.01 (-0.01; 0.04) | $2.2\times10^{-1}$ | <0.001 | 461 | IVW |
| | | Alzheimer | 0.97 (0.84; 1.12) | $6.7\times10^{-1}$ | 0.073 | 512 | IVW |
| | | Alzheimer, late onset | 0.51 (0.06; 4.47) | $5.5\times10^{-1}$ | 0.068 | 509 | MR Egger |
| | | Lewy body dementia | 0.60 (0.06; 5.55) | $6.5\times10^{-1}$ | 0.566 | 426 | IVW |
| LA – V (max) | SD | Any stroke | 1.14 (1.00; 1.29) | $4.3\times10^{-2}$ | 0.098 | 20 | IVW |
| | | Any ischemic stroke | 1.14 (1.01; 1.30) | $4.1\times10^{-2}$ | 0.218 | 20 | IVW |
| | | Large artery stroke | 1.22 (0.86; 1.73) | $2.7\times10^{-1}$ | 0.065 | 20 | IVW |
| | | Cardioembolic stroke | 1.10 (0.88; 1.37) | $4.0\times10^{-1}$ | 0.020 | 20 | IVW |
| | | Small vessel stroke | 0.96 (0.70; 1.32) | $8.0\times10^{-1}$ | 0.077 | 20 | IVW |
| | | VTE | 0.87 (0.47; 1.60) | $6.4\times10^{-1}$ | 0.043 | 15 | MR Egger |
| | | AAA | 0.31 (0.10; 0.93) | $3.7\times10^{-2}$ | 0.042 | 19 | MR Egger |
| | | SBP | -8.90 (-11.92; -5.88) | $7.6\times10^{-9}$ | 0.004 | 7 | MR Egger |
| | | DBP | -7.85 (-10.86; -4.84) | $3.1\times10^{-7}$ | 0.089 | 6 | MR Egger |
| | | BMI | -0.02 (-0.15; 0.11) | $7.8\times10^{-1}$ | <0.001 | 9 | MR Egger |
| | | T2DM | 1.23 (1.11; 1.36) | $4.9\times10^{-5}$ | 0.002 | 12 | IVW |
| | | Glycated haemoglobin | -2.53 (-3.87; -1.19) | $2.2\times10^{-4}$ | <0.001 | 13 | MR Egger |
| | | CRP | -0.56 (-0.86; -0.25) | $3.6\times10^{-4}$ | <0.001 | 14 | MR Egger |
| | | FEV1 | -0.11 (-0.20; -0.02) | $1.8\times10^{-2}$ | 0.086 | 19 | IVW |
| | | FVC | -0.07 (-0.15; 0.02) | $1.3\times10^{-1}$ | 0.164 | 19 | IVW |
| | | CKD | 1.00 (0.90; 1.12) | $9.6\times10^{-1}$ | 0.007 | 19 | IVW |
| | | eGFR | -0.07 (-0.10; -0.04) | $1.6\times10^{-5}$ | 0.057 | 13 | MR Egger |
| | | Alzheimer | 0.98 (0.95; 1.02) | $3.2\times10^{-1}$ | 0.079 | 20 | IVW |
| | | Alzheimer, late onset | 0.82 (0.70; 0.96) | $1.6\times10^{-2}$ | 0.021 | 20 | IVW |
| | | Lewy body dementia | 0.77 (0.50; 1.19) | $2.5\times10^{-1}$ | 0.373 | 19 | IVW |
| LA – V (min) | SD | Any stroke | 1.09 (0.91; 1.30) | $3.4\times10^{-1}$ | 0.248 | 11 | IVW |
| | | Any ischemic stroke | 1.01 (0.86; 1.20) | $8.7\times10^{-1}$ | 0.791 | 11 | IVW |
| | | Large artery stroke | 0.00 (0.00; 7.51) | $8.8\times10^{-2}$ | 0.032 | 10 | MR Egger |
| | | Cardioembolic stroke | 0.00 (0.00; 5.83) | $8.8\times10^{-2}$ | 0.238 | 10 | MR Egger |
| | | Small vessel stroke | 0.67 (0.42; 1.06) | $8.5\times10^{-2}$ | 0.161 | 11 | IVW |
| | | VTE | 0.99 (0.82; 1.20) | $9.2\times10^{-1}$ | 0.130 | 10 | IVW |
| | | AAA | 1.19 (0.90; 1.57) | $2.1\times10^{-1}$ | 0.742 | 11 | IVW |
| | | SBP | -0.09 (-0.63; 0.45) | $7.4\times10^{-1}$ | 0.005 | 9 | IVW |
| | | DBP | -0.02 (-0.33; 0.28) | $8.9\times10^{-1}$ | 0.026 | 10 | IVW |
| | | BMI | -0.01 (-0.04; 0.02) | $4.0\times10^{-1}$ | 0.017 | 10 | IVW |

**Supplementary Table 2:** Mendelian randomization phewas results of the effects of CMR measured changes in funtion and structure have on disease traits. *(continued)*

| CMR measure | Units | Cardiac diagnosis | OR (95%CI) | P-value | Q p-value | No. variants | MR model |
|---|---|---|---|---|---|---|---|
| | | T2DM | 1.21 (0.01; 108.89) | $9.3\times10^{-1}$ | 0.469 | 7 | MR Egger |
| | | Glycated haemoglobin | -0.35 (-0.63; -0.07) | $1.4\times10^{-2}$ | 0.261 | 11 | IVW |
| | | CRP | 2.96 (0.18; 5.75) | $3.7\times10^{-2}$ | 0.470 | 9 | MR Egger |
| | | FEV1 | -0.08 (-0.18; 0.02) | $1.0\times10^{-1}$ | 0.024 | 12 | IVW |
| | | FVC | 0.47 (-0.26; 1.19) | $2.1\times10^{-1}$ | 0.084 | 11 | MR Egger |
| | | CKD | 1.18 (0.97; 1.44) | $9.2\times10^{-2}$ | 0.142 | 11 | IVW |
| | | eGFR | -0.01 (-0.01; -0.00) | $1.1\times10^{-2}$ | 0.037 | 10 | IVW |
| | | Alzheimer | 1.04 (1.00; 1.09) | $6.5\times10^{-2}$ | 0.236 | 11 | IVW |
| | | Alzheimer, late onset | 0.62 (0.12; 3.23) | $5.7\times10^{-1}$ | 0.068 | 10 | MR Egger |
| | | Lewy body dementia | 1.15 (0.47; 2.81) | $7.5\times10^{-1}$ | 0.052 | 10 | IVW |
| LA – TF | SD | Any stroke | 0.88 (0.43; 1.80) | $7.2\times10^{-1}$ | 0.619 | 11 | MR Egger |
| | | Any ischemic stroke | 0.68 (0.32; 1.46) | $3.3\times10^{-1}$ | 0.450 | 12 | MR Egger |
| | | Large artery stroke | 8.49 (1.32; 54.85) | $2.5\times10^{-2}$ | 0.006 | 13 | MR Egger |
| | | Cardioembolic stroke | 0.57 (0.43; 0.77) | $1.6\times10^{-4}$ | 0.148 | 14 | IVW |
| | | Small vessel stroke | 0.85 (0.60; 1.19) | $3.4\times10^{-1}$ | 0.152 | 14 | IVW |
| | | VTE | 1.15 (1.01; 1.31) | $2.9\times10^{-2}$ | 0.636 | 13 | IVW |
| | | AAA | 1.25 (0.36; 4.28) | $7.3\times10^{-1}$ | 0.912 | 11 | MR Egger |
| | | SBP | 10.63 (2.33; 18.93) | $1.2\times10^{-2}$ | 0.005 | 6 | MR Egger |
| | | DBP | 2.82 (-1.71; 7.35) | $2.2\times10^{-1}$ | 0.982 | 7 | MR Egger |
| | | BMI | -0.00 (-0.03; 0.03) | $9.8\times10^{-1}$ | 0.093 | 13 | IVW |
| | | T2DM | 1.15 (1.05; 1.26) | $3.0\times10^{-3}$ | 0.016 | 12 | IVW |
| | | Glycated haemoglobin | -1.06 (-2.08; -0.04) | $4.2\times10^{-2}$ | 0.003 | 12 | MR Egger |
| | | CRP | -0.04 (-0.10; 0.03) | $2.8\times10^{-1}$ | 0.160 | 12 | IVW |
| | | FEV1 | 0.03 (-0.07; 0.13) | $5.6\times10^{-1}$ | 0.195 | 14 | IVW |
| | | FVC | 0.03 (-0.06; 0.12) | $5.4\times10^{-1}$ | 0.496 | 14 | IVW |
| | | CKD | 1.68 (0.56; 5.03) | $3.5\times10^{-1}$ | 0.335 | 11 | MR Egger |
| | | eGFR | 0.01 (0.00; 0.01) | $1.5\times10^{-3}$ | 0.105 | 13 | IVW |
| | | Alzheimer | 1.27 (0.96; 1.68) | $9.8\times10^{-2}$ | 0.830 | 13 | MR Egger |
| | | Alzheimer, late onset | 0.74 (0.60; 0.90) | $3.1\times10^{-3}$ | 0.567 | 13 | IVW |
| | | Lewy body dementia | 3.60 (0.05; 258.52) | $5.6\times10^{-1}$ | 0.336 | 11 | MR Egger |
| LA – AF | SD | Any stroke | 0.88 (0.76; 1.02) | $9.0\times10^{-2}$ | 0.153 | 20 | IVW |
| | | Any ischemic stroke | 0.92 (0.80; 1.06) | $2.4\times10^{-1}$ | 0.026 | 20 | IVW |
| | | Large artery stroke | 0.86 (0.59; 1.26) | $4.4\times10^{-1}$ | 0.204 | 20 | IVW |
| | | Cardioembolic stroke | 0.88 (0.67; 1.16) | $3.7\times10^{-1}$ | 0.041 | 19 | IVW |
| | | Small vessel stroke | 1.01 (0.70; 1.45) | $9.5\times10^{-1}$ | 0.170 | 20 | IVW |
| | | VTE | 1.07 (0.96; 1.19) | $2.5\times10^{-1}$ | 0.482 | 21 | IVW |
| | | AAA | 1.00 (0.79; 1.26) | $1.0\times10^{0}$ | 0.609 | 20 | IVW |
| | | SBP | 0.18 (-0.23; 0.60) | $3.9\times10^{-1}$ | 0.002 | 17 | IVW |
| | | DBP | -0.04 (-0.27; 0.20) | $7.5\times10^{-1}$ | 0.004 | 19 | IVW |
| | | BMI | -0.02 (-0.05; 0.01) | $1.3\times10^{-1}$ | 0.096 | 18 | IVW |
| | | T2DM | 1.11 (1.02; 1.20) | $1.7\times10^{-2}$ | 0.014 | 19 | IVW |
| | | Glycated haemoglobin | 0.24 (0.04; 0.44) | $1.6\times10^{-2}$ | <0.001 | 19 | IVW |
| | | CRP | 0.01 (-0.05; 0.06) | $8.6\times10^{-1}$ | 0.505 | 19 | IVW |
| | | FEV1 | 0.05 (-0.04; 0.13) | $2.6\times10^{-1}$ | 0.442 | 20 | IVW |
| | | FVC | 0.07 (-0.02; 0.17) | $1.3\times10^{-1}$ | 0.224 | 20 | IVW |
| | | CKD | 0.87 (0.75; 1.02) | $8.1\times10^{-2}$ | 0.126 | 20 | IVW |
| | | eGFR | 0.01 (0.01; 0.02) | $4.3\times10^{-5}$ | 0.347 | 20 | IVW |
| | | Alzheimer | 0.96 (0.93; 0.98) | $2.5\times10^{-3}$ | 0.298 | 20 | IVW |
| | | Alzheimer, late onset | 0.67 (0.54; 0.82) | $1.5\times10^{-4}$ | 0.346 | 19 | IVW |
| | | Lewy body dementia | 0.87 (0.49; 1.55) | $6.4\times10^{-1}$ | 0.125 | 19 | IVW |
| LA – PF | SD | Any stroke | 0.76 (0.62; 0.92) | $5.6\times10^{-3}$ | 0.542 | 7 | IVW |
| | | Any ischemic stroke | 0.78 (0.63; 0.96) | $1.9\times10^{-2}$ | 0.567 | 7 | IVW |
| | | Large artery stroke | 0.86 (0.50; 1.46) | $5.7\times10^{-1}$ | 0.927 | 7 | IVW |
| | | Cardioembolic stroke | 0.53 (0.34; 0.83) | $5.7\times10^{-3}$ | 0.285 | 7 | IVW |
| | | Small vessel stroke | 0.71 (0.41; 1.22) | $2.1\times10^{-1}$ | 0.291 | 7 | IVW |
| | | VTE | 1.09 (0.88; 1.36) | $4.4\times10^{-1}$ | 0.289 | 7 | IVW |
| | | AAA | 1.03 (0.71; 1.49) | $8.8\times10^{-1}$ | 0.345 | 7 | IVW |
| | | SBP | 0.39 (-0.29; 1.06) | $2.6\times10^{-1}$ | 0.757 | 6 | IVW |
| | | DBP | -0.53 (-2.15; 1.09) | $5.2\times10^{-1}$ | 0.086 | 7 | MR Egger |
| | | BMI | 0.02 (-0.03; 0.06) | $4.6\times10^{-1}$ | 0.191 | 7 | IVW |
| | | T2DM | 0.90 (0.79; 1.02) | $1.0\times10^{-1}$ | 0.440 | 7 | IVW |
| | | Glycated haemoglobin | -0.25 (-0.60; 0.09) | $1.5\times10^{-1}$ | 0.147 | 9 | IVW |
| | | CRP | -0.00 (-0.09; 0.09) | $9.9\times10^{-1}$ | 0.670 | 7 | IVW |
| | | FEV1 | 0.13 (-0.00; 0.26) | $5.1\times10^{-2}$ | 0.849 | 7 | IVW |
| | | FVC | 0.09 (-0.04; 0.23) | $1.6\times10^{-1}$ | 0.995 | 7 | IVW |
| | | CKD | 0.82 (0.68; 1.00) | $5.4\times10^{-2}$ | 0.607 | 7 | IVW |

**Supplementary Table 2:** Mendelian randomization phewas results of the effects of CMR measured changes in funtion and structure have on disease traits. *(continued)*

| CMR measure | Units | Cardiac diagnosis | OR (95%CI) | P-value | Q p-value | No. variants | MR model |
|---|---|---|---|---|---|---|---|
| | | eGFR | 0.00 (-0.01; 0.01) | $4.1\times10^{-1}$ | 0.110 | 7 | IVW |
| | | Alzheimer | 0.98 (0.94; 1.02) | $3.8\times10^{-1}$ | 0.970 | 7 | IVW |
| | | Alzheimer, late onset | 0.91 (0.68; 1.23) | $5.4\times10^{-1}$ | 0.018 | 7 | IVW |
| | | Lewy body dementia | 1.12 (0.53; 2.35) | $7.7\times10^{-1}$ | 0.500 | 7 | IVW |
| LA – PFR | 20 ml/s | Any stroke | 0.99 (0.92; 1.07) | $7.7\times10^{-1}$ | 0.234 | 2 | IVW |
| | | Any ischemic stroke | 0.98 (0.92; 1.05) | $5.6\times10^{-1}$ | 0.334 | 2 | IVW |
| | | Large artery stroke | 6.25 (1.59; 24.48) | $8.6\times10^{-3}$ | None | 2 | MR Egger |
| | | Cardioembolic stroke | 0.92 (0.81; 1.05) | $2.1\times10^{-1}$ | 0.285 | 2 | IVW |
| | | Small vessel stroke | 1.09 (0.94; 1.27) | $2.5\times10^{-1}$ | 0.484 | 2 | IVW |
| | | VTE | 0.95 (0.86; 1.06) | $3.8\times10^{-1}$ | 0.790 | 2 | IVW |
| | | AAA | 0.99 (0.80; 1.22) | $9.1\times10^{-1}$ | 0.192 | 2 | IVW |
| | | SBP | | | | | |
| | | DBP | 0.31 (0.13; 0.50) | $9.9\times10^{-4}$ | None | 1 | Wald |
| | | BMI | -0.01 (-0.03; 0.00) | $1.4\times10^{-1}$ | None | 1 | Wald |
| | | T2DM | 1.12 (1.06; 1.19) | $7.0\times10^{-5}$ | 0.139 | 2 | IVW |
| | | Glycated haemoglobin | 0.16 (-0.11; 0.43) | $2.5\times10^{-1}$ | None | 2 | MR Egger |
| | | CRP | | | | | |
| | | FEV1 | -0.02 (-0.05; 0.01) | $2.6\times10^{-1}$ | 0.008 | 3 | IVW |
| | | FVC | -0.01 (-0.04; 0.03) | $7.2\times10^{-1}$ | 0.225 | 3 | IVW |
| | | CKD | 1.02 (0.91; 1.13) | $7.7\times10^{-1}$ | None | 1 | Wald |
| | | eGFR | 0.01 (0.00; 0.01) | $3.5\times10^{-3}$ | None | 1 | Wald |
| | | Alzheimer | 1.01 (0.96; 1.07) | $6.3\times10^{-1}$ | 0.841 | 2 | IVW |
| | | Alzheimer, late onset | 0.90 (0.84; 0.97) | $3.4\times10^{-3}$ | 0.556 | 3 | IVW |
| | | Lewy body dementia | 0.76 (0.52; 1.11) | $1.5\times10^{-1}$ | None | 1 | Wald |
| RA – PFR | 20 ml/s | Any stroke | 1.02 (0.95; 1.09) | $6.5\times10^{-1}$ | 0.140 | 4 | IVW |
| | | Any ischemic stroke | 1.00 (0.93; 1.07) | $9.0\times10^{-1}$ | 0.172 | 4 | IVW |
| | | Large artery stroke | 0.99 (0.82; 1.19) | $8.9\times10^{-1}$ | 0.125 | 5 | IVW |
| | | Cardioembolic stroke | 0.95 (0.85; 1.06) | $3.7\times10^{-1}$ | 0.385 | 4 | IVW |
| | | Small vessel stroke | 1.05 (0.90; 1.23) | $5.5\times10^{-1}$ | 0.208 | 4 | IVW |
| | | VTE | 0.89 (0.81; 0.98) | $1.5\times10^{-2}$ | 0.528 | 5 | MR Egger |
| | | AAA | 0.80 (0.67; 0.95) | $9.9\times10^{-3}$ | 0.981 | 5 | MR Egger |
| | | SBP | 0.11 (-0.06; 0.27) | $2.0\times10^{-1}$ | 0.697 | 4 | IVW |
| | | DBP | 0.11 (0.01; 0.21) | $3.0\times10^{-2}$ | 0.221 | 5 | IVW |
| | | BMI | -0.00 (-0.01; 0.01) | $9.8\times10^{-1}$ | 0.135 | 5 | IVW |
| | | T2DM | 1.02 (0.99; 1.06) | $1.2\times10^{-1}$ | 0.648 | 5 | IVW |
| | | Glycated haemoglobin | 0.03 (-0.03; 0.09) | $3.1\times10^{-1}$ | 0.495 | 7 | IVW |
| | | CRP | 0.00 (-0.02; 0.02) | $9.9\times10^{-1}$ | 0.255 | 5 | IVW |
| | | FEV1 | -0.02 (-0.04; 0.01) | $2.9\times10^{-1}$ | 0.395 | 6 | IVW |
| | | FVC | -0.02 (-0.05; 0.01) | $1.5\times10^{-1}$ | 0.298 | 6 | IVW |
| | | CKD | 1.04 (0.99; 1.09) | $1.1\times10^{-1}$ | 0.045 | 5 | IVW |
| | | eGFR | -0.00 (-0.00; 0.00) | $5.5\times10^{-2}$ | 0.648 | 5 | IVW |
| | | Alzheimer | 1.00 (0.99; 1.02) | $3.8\times10^{-1}$ | 0.424 | 5 | IVW |
| | | Alzheimer, late onset | 1.04 (0.94; 1.14) | $4.5\times10^{-1}$ | 0.129 | 5 | IVW |
| | | Lewy body dementia | 0.83 (0.68; 1.01) | $5.7\times10^{-2}$ | 0.696 | 4 | IVW |

*General:*
CMR: Cardiac MRI, MR: Mendelian randomization, OR: odds ratio difference, CI: confidence interval, Q: Q test for heterogeneity. Effect estimates are coded towards the CMR increasing direction.

**Supplementary Table 3:** Mendelian randomization phewas results of the effects Atrial fibrillation and Heart failure have on disease traits.

| CMR measure | Cardiac diagnosis | OR (95%CI) | P-value | Q p-value | No. variants | MR model |
|---|---|---|---|---|---|---|
| Atrial fibrillation | Any stroke | 1.25 (0.99; 1.56) | $5.8\times10^{-2}$ | 0.407 | 151 | MR Egger |
| | Any ischemic stroke | 1.15 (0.89; 1.48) | $2.9\times10^{-1}$ | 0.181 | 152 | MR Egger |
| | Large artery stroke | 1.11 (0.99; 1.24) | $8.0\times10^{-2}$ | 0.007 | 161 | IVW |
| | Cardioembolic stroke | 2.13 (1.35; 3.34) | $1.1\times10^{-3}$ | 0.032 | 153 | MR Egger |
| | Small vessel stroke | 0.99 (0.57; 1.74) | $9.8\times10^{-1}$ | 0.019 | 152 | MR Egger |
| | VTE | 1.03 (0.82; 1.30) | $8.0\times10^{-1}$ | <0.001 | 147 | MR Egger |
| | AAA | 0.89 (0.62; 1.29) | $5.5\times10^{-1}$ | 0.033 | 152 | MR Egger |
| | SBP | -0.64 (-1.55; 0.26) | $1.6\times10^{-1}$ | <0.001 | 113 | MR Egger |
| | DBP | -0.95 (-1.49; -0.40) | $6.6\times10^{-4}$ | <0.001 | 108 | MR Egger |
| | BMI | -0.03 (-0.08; 0.02) | $2.6\times10^{-1}$ | <0.001 | 120 | MR Egger |
| | T2DM | 1.12 (0.95; 1.33) | $1.8\times10^{-1}$ | <0.001 | 136 | MR Egger |
| | Glycated haemoglobin | 0.00 (-0.07; 0.08) | $9.0\times10^{-1}$ | <0.001 | 150 | IVW |
| | CRP | -0.02 (-0.04; -0.00) | $3.1\times10^{-2}$ | <0.001 | 156 | IVW |
| | FEV1 | 0.02 (-0.13; 0.17) | $7.8\times10^{-1}$ | <0.001 | 145 | MR Egger |
| | FVC | 0.01 (-0.02; 0.04) | $3.4\times10^{-1}$ | <0.001 | 156 | IVW |
| | CKD | 0.95 (0.76; 1.18) | $6.3\times10^{-1}$ | <0.001 | 151 | MR Egger |
| | eGFR | 0.01 (-0.00; 0.02) | $8.6\times10^{-2}$ | <0.001 | 142 | MR Egger |
| | Alzheimer | 0.98 (0.93; 1.03) | $3.9\times10^{-1}$ | 0.036 | 158 | MR Egger |
| | Alzheimer, late onset | 0.81 (0.58; 1.13) | $2.2\times10^{-1}$ | 0.666 | 154 | MR Egger |
| | Lewy body dementia | 1.37 (0.51; 3.72) | $5.3\times10^{-1}$ | 0.017 | 127 | MR Egger |
| Heart failure | Any stroke | 1.47 (1.33; 1.63) | $6.6\times10^{-14}$ | 0.005 | 26 | IVW |
| | Any ischemic stroke | 1.50 (1.35; 1.67) | $2.3\times10^{-13}$ | 0.002 | 27 | IVW |
| | Large artery stroke | 2.10 (1.62; 2.71) | $1.7\times10^{-8}$ | 0.039 | 29 | IVW |
| | Cardioembolic stroke | 1.49 (0.76; 2.95) | $2.5\times10^{-1}$ | 0.025 | 24 | MR Egger |
| | Small vessel stroke | 1.31 (1.00; 1.70) | $5.0\times10^{-2}$ | 0.214 | 28 | IVW |
| | VTE | 1.14 (1.04; 1.25) | $4.8\times10^{-3}$ | 0.017 | 35 | IVW |
| | AAA | 1.31 (1.10; 1.54) | $1.9\times10^{-3}$ | 0.136 | 37 | IVW |
| | SBP | 0.93 (0.56; 1.30) | $7.0\times10^{-7}$ | <0.001 | 18 | IVW |
| | DBP | 0.32 (0.04; 0.61) | $2.6\times10^{-2}$ | 0.158 | 15 | IVW |
| | BMI | -0.04 (-0.09; 0.01) | $1.2\times10^{-1}$ | 0.003 | 20 | MR Egger |
| | T2DM | 0.78 (0.66; 0.91) | $2.0\times10^{-3}$ | <0.001 | 27 | MR Egger |
| | Glycated haemoglobin | 0.06 (-0.09; 0.20) | $4.5\times10^{-1}$ | <0.001 | 31 | IVW |
| | CRP | 0.05 (0.00; 0.10) | $4.7\times10^{-2}$ | 0.121 | 28 | IVW |
| | FEV1 | -0.06 (-0.11; -0.00) | $4.7\times10^{-2}$ | 0.027 | 36 | IVW |
| | FVC | -0.04 (-0.10; 0.01) | $1.4\times10^{-1}$ | <0.001 | 36 | IVW |
| | CKD | 1.25 (1.14; 1.37) | $4.5\times10^{-6}$ | 0.014 | 29 | IVW |
| | eGFR | -0.01 (-0.01; -0.00) | $9.2\times10^{-5}$ | 0.003 | 28 | IVW |
| | Alzheimer | 1.01 (0.98; 1.03) | $5.8\times10^{-1}$ | 0.815 | 32 | IVW |
| | Alzheimer, late onset | 0.91 (0.80; 1.04) | $1.8\times10^{-1}$ | 0.401 | 35 | IVW |
| | Lewy body dementia | 1.22 (0.83; 1.78) | $3.2\times10^{-1}$ | 0.140 | 31 | IVW |

*General:*
MR: Mendelian randomization, OR: odds ratio difference, CI: confidence interval, Q: Q test for heterogeneity.
